# Ulcerative Colitis Host-Microbiome Response to Hyperbaric Oxygen Therapy

**DOI:** 10.1101/2022.01.14.22269325

**Authors:** Carlos G. Gonzalez, Robert H. Mills, Melissa C. Kordahi, Marvic Carrillo-Terrazas, Henrry Secaira-Morocho, Christella E. Widjaja, Matthew S. Tsai, Yash Mittal, Brian A. Yee, Fernando Vargas, Kelly Weldon, Julia M. Gauglitz, Clara Delaroque, Consuelo Sauceda, Leigh-Ana Rossitto, Gail Ackermann, Gregory Humphrey, Austin D. Swafford, Corey A. Siegel, Jay C. Buckey, Laura E. Raffals, Charlotte Sadler, Peter Lindholm, Kathleen M. Fisch, Mark Valaseck, Arief Suriawinata, Gene W. Yeo, Pradipta Ghosh, John T. Chang, Hiutung Chu, Pieter Dorrestein, Qiyun Zhu, Benoit Chassaing, Rob Knight, David J. Gonzalez, Parambir S. Dulai

**Author notes:** **Correspondence:** Parambir S. Dulai, MD, Associate Professor of Medicine, Director, GI Clinical Trials, Director, Precision Medicine, Division of Gastroenterology & Hepatology, Northwestern University Feinberg School of Medicine, Chicago, IL 60611. **Author Contributions:** Cohort recruitment and sample collection: PSD, CAS, JCB, LER, CS, PL; Data acquisition and/or preparation: CGG, RHM, MK, MVCT, HSM, CEW, BAY, FV, KW, JMG, CD, CS, LR, GA, GH, KMF, MV, AS, YM, PSD, CS, LAR; Data analysis: CGG, RHM, MK, HSM, BAY, ADS, KMF, GWY, PG, JTC, HC, PD, RK, DJG, PSD; Manuscript drafting: CGG, RHM, MST, CAS, JCB, LER, KMF, PG, JTC, HC, PD, RK, DJG, PSD; study oversight: PSD, DJG, RK, PD, HC, JTC, PG, GWY, BC, QZ. Critical revisions and approval of final manuscript: all authors. **Disclosures:** None related to the work being presented.

## Abstract

**Objective:** This study examined the host-microbe changes underpinning treatment response to hyperbaric oxygen therapy (HBOT) in ulcerative colitis.

**Design:** Pre- and post-intervention mucosal tissue and fecal samples from two clinical trials, along with fecal samples from healthy controls and fecal and mucosal tissue from disease severity matched UC controls. Mucosal tissue bulk-RNA sequencing, digital spatial profiling (DSP) for single-cell RNA and protein analysis, and immunohistochemistry was performed, in addition to 16S rRNA, shotgun metagenomics, metabolomics, and metaproteomics of fecal samples. Fecal colonization experiments in IL10^-/-^ germ-free mice were performed to confirm observations.

**Results:** Proteomics identified associations between HBOT response status and neutrophil degranulation, with specificity of effect for azurophilic granules. DSP identified a specific HBOT effect on reducing neutrophil STAT3, which was confirmed by immunohistochemistry. HBOT decreased microbial diversity with an accompanying proportional increase in Firmicutes and a secondary bile acid lithocholic acid. The reduction in diversity was due to reductions in mucinophiles, with differences in *Akkermansia muciniphila* strains being associated with HBOT response status. Proteomics observed an accompanying effect for HBOT on MUC2. Colonization of IL10^-/-^ with stool obtained from HBOT responders resulted in lower colitis activity compared to stool obtained from HBOT non-responders, with no differences in STAT3 expression, suggesting complementary but independent host and microbial responses.

**Conclusion:** HBOT reduces host neutrophil STAT3 and azurophilic granule activity in UC patients, and changes in microbial composition and metabolism in ways that improve colitis activity. Intestinal microbiota, especially strain level variations in *A. muciniphila*, may contribute to HBOT non-response.

## INTRODUCTION

Maintenance of oxygen homeostasis is essential to survival, and the human body has developed adaptative programs of transcriptional response to hypoxia which allow for maintenance. Dysregulation of these responses is a core feature of ulcerative colitis (UC), where tissue hypoxia results in epithelial barrier disruption, microbial dysbiosis, recruitment and activation of neutrophils and pathogenic T-cells, and overproduction of inflammatory cytokines.^1^ These changes further augment microenvironmental hypoxia and ultimately lead to worsening colonic inflammation.^1^ Therapeutics are in development targeting aspects of these dysregulated hypoxia response pathways.^2^ However, it remains to be defined what the key mediators of this complex host-microbe response are.

In a phase 2A multi-center, double-blind, sham-controlled, randomized trial for moderate-severe hospitalized UC patients, we demonstrated that the intermittent delivery of high amounts of oxygen to the colon through hyperbaric oxygen therapy (HBOT) resulted in significantly greater rates of clinical response and remission, and significantly lower rates of progression to infliximab or colectomy during the hospitalization.^3^ In a subsequent phase 2B multi-center, dose-finding, randomized trial, we further confirmed treatment efficacy of HBOT in hospitalized UC patients and observed 5 HBOT sessions to be superior to 3 HBOT sessions for improving disease outcomes.^4^ In the current study we aimed to define the host-microbe response to HBOT in UC to better understand mechanisms through which targeting hypoxia may result in disease remission. Insights gained from our work will be of importance as we target hypoxia for the treatment of UC and other inflammatory disorders. The multi-omics approach and digital spatial profiling (DSP) performed will be of value to researchers looking to apply these technologies to other diseases, particularly when attempting to study the role of neutrophils in intestinal inflammation given technical challenges and complexities with isolating these immune cells from mucosal tissue.

## METHODS

### Clinical Trial Overview

We utilized clinical data and human biospecimens collected from the phase 2 clinical trials,^3,4^ for HBOT in UC patients hospitalized for a medically refractory moderate to severe UC flare (Full Mayo score ≥ 6, all sub-scores 2-3). All patients received intravenous steroids along with either HBOT (2.4 atmospheres absolute [ATA; sea level], 100% oxygen, 90-minute sessions, one session per day) or sham air (1.34 ATA, 21% oxygen, 90-minute sessions, one session per day), over 3-5 days. Disease activity was measured using the Mayo score. Endoscopy sub-scores were available at study days 0 and 10, with rectal bleeding and stool frequency sub-scores available at days 0, 3, 5, and 10. The institutional review board at each center approved the protocols, and all patients gave written informed consent for participation in the trial and the translational research outlined using human bio specimens collected. Trial details can be found in the original publications.^3,4^ Trial registration can be found at clinicaltrials.gov (#NCT02144350).

### Mucosal Tissue RNA and protein profiling

Bulk-RNA sequencing was done for available pre- and post-intervention mucosal biopsies (n=5 HBOT and n=8 sham treated patients) from the sham-controlled phase 2A trial based on standard pipelines at the University of California San Diego Institute for Genomic Medicine facility. (see **Supplementary Methods Bulk-RNA Sequencing**) DSP was done for pre- and post-intervention mucosal biopsies from 3 hospitalized UC patients treated with HBOT in the phase 2 trials, and 3 disease-severity and response-status matched outpatient UC controls not treated with HBOT. All 3 of the HBOT treated patients included in the DSP cohort had severe endoscopic activity (Mayo endoscopic sub-score 3) at day 0 with endoscopic improvement (Mayo endoscopic sub-score 0-1) at day 10. The 3 outpatient UC controls were matched to the 3 HBOT UC cases for baseline disease severity, biologic exposure status, and response status, with all 3 having severe endoscopic activity (Mayo endoscopic sub-score 3) at baseline and endoscopic improvement (Mayo endoscopic sub-score 0-1) at follow-up. The GeoMx DSP developed by NanoString Technologies enables spatially resolved, high-plex (10s -10,000s) digital quantitation of proteins and mRNA in tissue, and a pre-defined panel of mRNA and proteins was used. (**Supplementary Table 1**) DSP protocols were followed according to the manufacturer’s instructions and in accordance with prior published work using this platform.^5,6^ (see **Supplementary Methods Digital Spatial Profiling**) Mucosal proteomics was done for pre- and post-intervention mucosal biopsies from the phase 2B dose-finding trial (n=20 HBOT patients) based on standard pipelines at the University of California San Diego Collaborative Center of Multiplexing Proteomics, with comparison to mucosal proteomics datasets available publicly for UC (n=22) and non-IBD controls (n=24).^7^ (see **Supplementary Methods Proteomics**^8-10^).

### Immunohistochemistry

Immunohistochemistry (IHC) was performed for hypoxia inducible factor (HIF)-1α and heme-oxygenase (HO-1) with an *a priori* hypothesis that HBOT response status would correlate to changes in these hypoxia response pathways. Post-hoc IHC was done for STAT3 and phosphorylated STAT3 to confirm observations made through DSP and proteomics. (see **Supplementary Methods Immunohistochemistry**)

### Fecal Multi-Omics Experiments

Fecal samples from 43 participants (n=8 sham and n=25 HBOT from phase 2 trials, n=16 outpatient UC severity matched controls (IRB#150675), n=4 healthy controls recruited through the American Gut Project^11^) underwent 16S rRNA gene amplicon sequencing, shotgun metagenomics, metabolomics, and metaproteomics. Samples in the phase 2A sham-controlled trial were available at days 0 and 10, and samples from the phase 2B dose-finding trial were available from days 0, 3, 5, and 10. Analyses were adjusted for shifts in disease severity over time and matched against healthy controls and the outpatient UC cohort not treated with HBOT or sham air. In addition to our standard Qiime2-based shotgun metagenomic search pipeline, samples were further searched with a targeted database containing 279 strains of *A. muciniphila* in addition to the previous species-level entries found in our metagenomic database. (see **Supplementary Methods Fecal Proteomics, 16S gene amplicon sequencing, Metabolomics, and Metagenomics**^11-18^)

### Fecal Colonization Experiment

To confirm independence of effect for HBOT on host and microbe responses, and effects of microbial changes on host immune responses, we performed fecal colonization in germ-free IL10 knockout (IL10^-/-^) mice using post-intervention stool from patients treated with HBOT stratified by response status. At 6-8 weeks of age, mice were administered 100 μL of fecal suspension once and then transferred to micro-isolator cages and maintained with autoclaved food (Lab Diet). Two groups of mice (n=8-10 mice per group) were colonized, group 1 was colonized with stool collected from two UC patients who responded clinically to HBOT (at least a 2-point reduction in partial Mayo with at least 1 point reduction in both rectal bleeding and stool frequency sub-scores), and group 2 was colonized with stool collected from two UC patient who did not respond clinically to HBOT (0- or 1-point reduction in partial Mayo with no change in rectal bleeding sub-score). To ensure differences observed were not simply a function of mucosal remission status, the two HBOT responders chosen were patients who had begun to respond clinically to HBOT but who had persistent endoscopic inflammation (Mayo endoscopic sub-score 2) at the time of stool collection. This was to allow for an assessment of whether the evolving changes in microbial composition in these clinical responders were less colitogenic compared to the non-responders. (see **Supplementary Methods Fecal Colonization Experiments for Histology**^19^)

### Data Availability

Proteomic data and supplementary files are available online at https://massive.ucsd.edu (Study ID MSV000088636). Metaproteomic data used for the severity-matched patient analyses were derived from a previous study and is available at MassIVE (Study ID MSV000083874). Metabolomics files are available at massive.ucsd.edu (Study ID: MSV000081492 and MSV000086483). Processed 16s, shotgun metagenomic and metabolomic results are available online at qiita.ucsd.edu (Study ID: 11149). Bulk RNA sequencing data is available on NCBI server (accession ID: GSE152229). All results and scripts are available at https://github.com/c6gonzalez/HBOT and https://github.com/rhmills/HBOT_Multiomics. Patient demographics are reported in **Supplementary Tables 2 and 3**.

## RESULTS

### HBOT Stabilizes Mucosal Hypoxia Response Pathways Independently of Clinical Response Status

HBOT resulted in 1) increase in epithelial HIF-1α activity relative to sham (**figure 1a**), 2) a more uniform distribution in activity for stromal HO-1 relative to sham (**figure 1b**), and 3) an increase in gene expression for metallothionein (MT1 and 2) family members (figure **1c**). Proteomics of paired biopsies also revealed an increase in mucosal prostaglandin E2 (PGE2) synthase (**figure 1d)**. However, the effect of HBOT on these mucosal hypoxia response pathways was independent of response status and was observed in both HBOT responders and non-responders. **Supplementary Figures 1-3** provide representative IHC images, bulk-RNA sequencing results, and details for protein changes or inferences from mucosal and fecal datasets.

**Figure 1:**
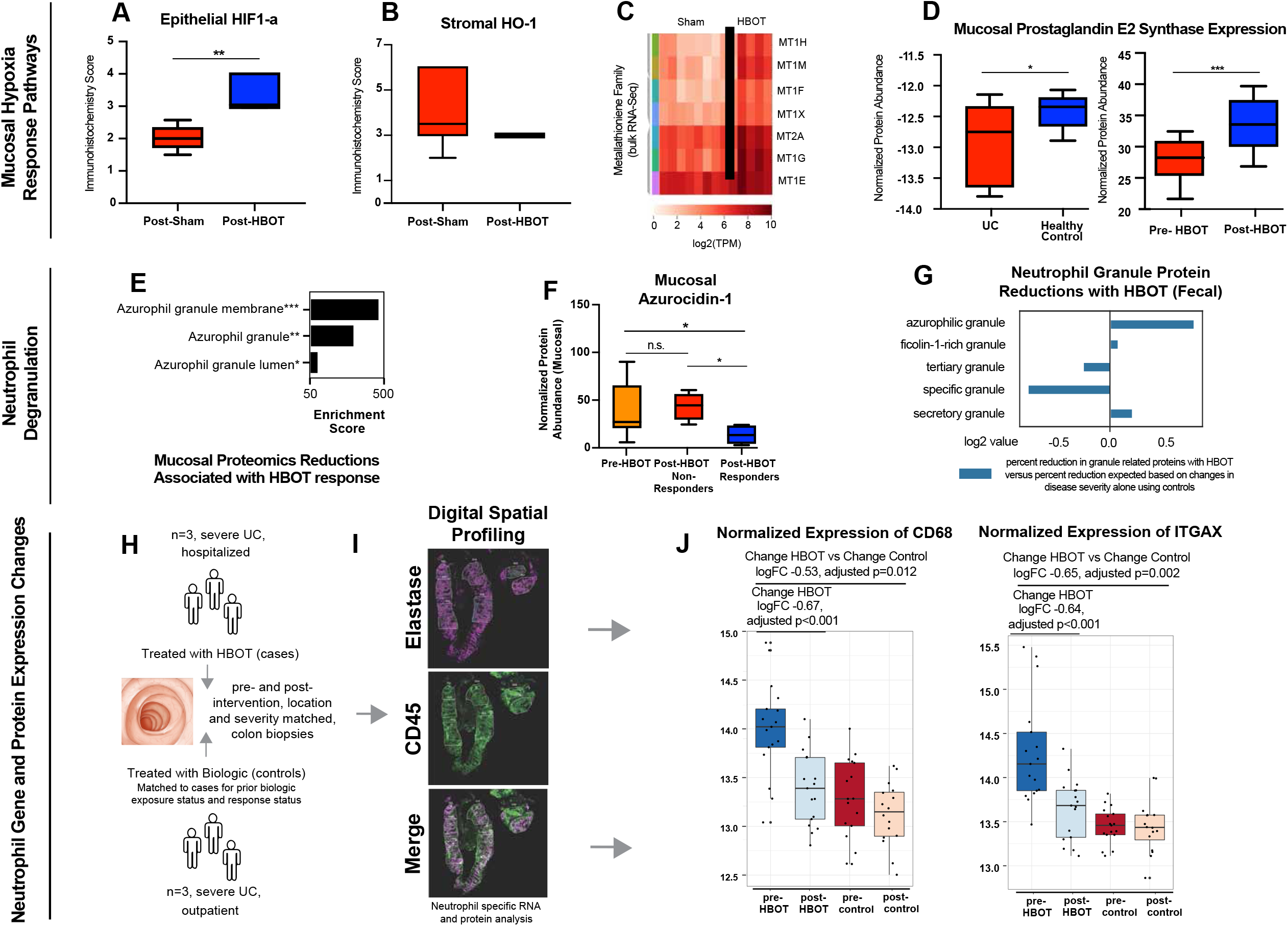
Translational Sequencing Studies Identify Effect for Hyperbaric Oxygen on Hypoxia Response Pathways and Neutrophil Degranulation. **A and B:** Boxplots of IHC score for epithelial HIF-1α and HO-1 activity for sham (n=8) and HBOT (n=10) treated UC patients. Upper and lower limits represent sample score range (A: p<0.001, unpaired t-test; B: not significant); **C:** Post-treatment Bulk-RNA sequencing results of mucosal biopsies for sham (n=8) and HBOT (n=5) treated UC patients enrolled in the phase 2A clinical trial. Metallothioneins are a family of highly conserved stress-induced proteins that, by control of cellular zinc homeostasis, protect against oxidative stress; **D:** Boxplot of PGE2 synthase proteome abundance in mucosal biopsies is demonstrated to be significantly lower in UC patients compared to healthy controls using a publicly available proteomics dataset (left bar graph, n=20, p = 0.03, unpaired t-test), and PGE2 synthase expression in mucosal biopsies is demonstrated to be significantly increased with HBOT exposure in the phase 2 trials (right bar graph; n=20, p < 0.0001, paired t-test). Ranges shown represent min. and max. sample measurement range. **E:** TMT-multiplexed proteomics-derived enrichments in proteins reduced by HBOT treatment were strongly associated in proteins associated with azurophilic granule membranes, granules, and granule lumen. Enrichment strength (plotted) is derived from the odds ratio and adjusted p-value, output from Enrichr algorithm. **F:** Day 10 pre- and post-HBOT mucosal proteome demonstrated a significant decrease in azurocidin-1 protein abundance in HBOT responders (n=5) compared to pre-HBOT (ANOVA-adjusted group-wise comparison p = 0.04) and non-responders (p=0.01), while non-responders were not significantly different from pre-HBOT condition (n=5, p>0.99); **G:** Pre- and post-HBOT fecal proteome datasets with severity matched UC controls demonstrates a specificity for effect of HBOT on azurophilic granules which was greater than would be expected with shifts in disease severity alone. **H and I:** Digital spatial profiling for neutrophils and other immune cells was done for mRNAs and proteins in 3 severe UC patients who were hospitalized and treated with HBOT compared to 3 severe UC patients who were managed in the outpatient setting with biologics. Cases and controls were matched for prior biologic exposure, number of prior biologics, and response status, and mucosal samples were matched for colon location and severity of activity pre- and post-intervention; **J:** Changes in neutrophil gene expression changes for CD68 and ITGAX with HBOT versus controls. CD68 is a membrane bound protein that is found in primary (azurophilic) neutrophil granules. *p<0.05; *p<0.01; ***p<0.001, ANOVA-adjusted. Range represents min and max sample measurements.

### HBOT Response Status Associated with Reductions in Azurophilic Granule Related Proteins

Proteomics of mucosal tissue and fecal samples both identified an enrichment of effect for HBOT on reducing neutrophil degranulation, with specificity of effect for reducing azurophilic granule related proteins (p<0.001 for both mucosal and fecal proteome datasets). This effect was more pronounced among HBOT responders compared to non-responders. (**figure 1e-g; supplementary figure 4** provides detailed neutrophil degranulation network map and protein changes with HBOT) In mucosal samples, a significant negative correlation was observed between response to HBOT and Azurocidin-1 (*r* = -0.65, p=0.04), and HBOT responders had significantly lower normalized protein abundances for Azurocidin-1 at day 10 follow-up compared to HBOT non-responders. (**figure 1f**) In fecal samples, reductions in azurophilic granule proteins observed with HBOT exposure were greater than would be expected with changes in disease activity alone, and this was observed in both HBOT clinical trial datasets independently. (**figure 1g**) Separate analyses linked proteins related to NOD-, LRR- and pyrin domain-containing protein 3 (NLRP3) with a positive response to HBOT. (**Supplementary figure 4e and f**).

To further probe neutrophil response, DSP on formalin fixed paraffin embedded (FFPE) mucosal tissue was then performed with a specific focus on neutrophils given observed effects of HBOT on neutrophil degranulation. (figure **1 h-j**) When comparing differences between groups (HBOT and controls) for within group (post-vs pre-) changes in neutrophil gene expression, a significant reduction in CD68 and ITGAX (Integrin alpha X) was observed with HBOT relative to controls. ITGAX is part of the neutrophil degranulation pathway, and CD68 is a membrane bound protein that is found in primary (azurophilic) neutrophil granules.^20^

### HBOT Reduces STAT3 Activity in Neutrophils

DSP of mRNA transcripts revealed an enrichment of STAT3 mediated interleukin signaling related to HBOT (p<0.001). HBOT treated patients had significantly reduced STAT3 and HIF-1α compared to controls treated with biologics. (**figure 2a and b**; **supplementary figures 5 and 6**) Each patient sample had between 5-7 neutrophil regions analyzed with DSP, allowing for an assessment of heterogeneity in effect and changes. HBOT resulted in a significant decrease in STAT3 across all neutrophil regions at follow-up for all treated patients, however this consistency in reduction was not observed for HIF-1α. (**supplementary figure 7**) Other notable neutrophil gene expression changes related to the STAT3 pathway are shown in **supplementary figure 8**. We observed no significant effect for HBOT on STAT3 or HIF-1α in the non-neutrophil immune cell regions (CD45^+^elastase^-^), (**supplementary figure 9**) demonstrating this effect of HBOT on STAT3 to be neutrophil specific.

**Figure 2:**
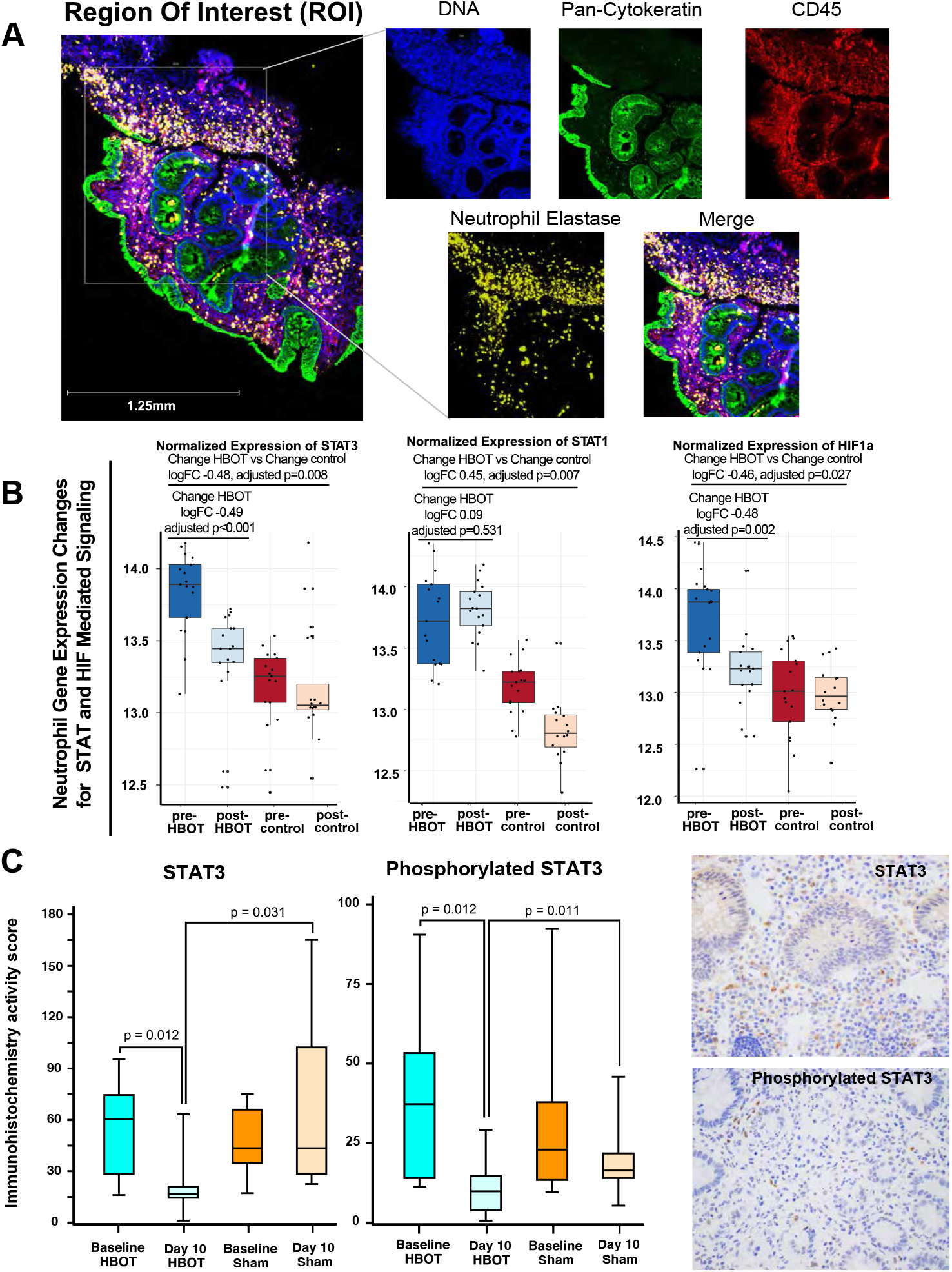
Hyperbaric Oxygen Therapy Reduces STAT3. **A:** Representative image for defining regions of interest (ROI) from mucosal tissue in patients with UC receiving HBOT (hyperbaric oxygen therapy), utilized for digital spatial profiling (Blue = DNA, Green = Pan-cytokeratin, Red = CD45, Yellow = Neutrophil elastase). **B:** Single-cell gene expression changes of STAT3 (left), STAT1 (center), HIF1a (right), in neutrophil-specific ROI (CD45+elastase+) in HBOT treated vs. matched control patients with UC not treated with HBOT. Each dot represents a unique ROI within a single sample, and on average 6 neutrophil specific ROIs were available per sample per patient. Ranges represent max and min calculated values. **C:** Boxplot of immunohistochemistry scores and staining for STAT3 and phosphorylated STAT3 done for mucosal biopsies. Measurements were taken pre- and post-exposure to sham (n=8) or HBOT (n=22) in the phase 2 trials. Ranges represent absolute min. and max. Sample values. Statistical significance derived using ANOVA-adjusted p-value calculations.

This effect of HBOT on STAT3 was confirmed using immunohistochemistry and observed that HBOT treated patients had a significant reduction in both STAT3 and phosphorylated STAT3 at study day 10 compared to baseline, with no significant changes in these markers in the sham treated patients, and day 10 values for STAT3 and phosphorylated STAT3 were significantly lower in HBOT versus sham treated patients from the randomized phase 2 clinical trials. (**figure 2c**)

### HBOT Reduces MAPK and Markers of Reactive Oxygen Species, and Increases the Pro-Apoptotic BIM and the NLRP3 Inhibitor BCLXL

In the neutrophil transcriptome datasets, a significant decrease was observed in CXCR6 with HBOT (logFC –0.61, adjusted p=0.002), known to be involved in ROS mediated injury via MAPK.^21^ In the neutrophil proteome datasets, HBOT treated patients had a significant reduction in protein abundance for phosphorylated p38 MAPK, phosphorylated ERK, and phosphorylated JNK relative to controls. (**supplementary figure 10**) In the proteome datasets, a significant effect for HBOT was observed for neutrophil apoptosis (p<0.001) and programmed cell death (p<0.01). Alongside the above noted neutrophil changes in MAPK and ROS mediated pathways, we observed a significant effect for HBOT on increasing neutrophil protein expression for B-cell lymphoma-extra large (BCLXL; logFC 0.69, adjusted p=0.008) and bcl-2 interacting protein (BIM; logFC 0.44, adjusted p=0.033). No significant effect was seen for HBOT on neutrophil protein expression for bcl-2-associated death promoter (BAD; adjusted p=0.62), and in the neutrophil transcriptome dataset no significant effect was seen for HBOT on neutrophil gene expression for bcl-2 (adjusted p=0.18).

### HBOT Reduces *Akkermansia* and Increases MUC2 in UC Patients

HBOT resulted in a relative increase in the Firmicutes:Bacteroidetes ratio within the metagenome and a reduction in host proteins relative to *Firmicutes* in the metaproteome. (**supplementary figure 11**) Fecal metabolomics observed an association between HBOT response status and lithocholic acid (LCA) levels (r=0.9 change in partial Mayo and LCA, p=0.0002). (**supplementary figure 12**) Within the 16S dataset, we observed this proportional increase in *Firmicutes* to be a result of a significant reduction in Shannon diversity with HBOT responders relative to sham treated patients. The degree of reduction in Shannon diversity was linearly related to degree of HBOT response and number of HBOT treatment sessions used. (**supplementary figure 11**)

The two most reduced families by HBOT were *Muribaculaceae* (formerly *Bacteroidales S24-7*) and *Akkermansiaceae*, (**figure 3a**) both known to be mucinophiles. Reductions in *A. muciniphila* specifically were significantly associated with reductions in partial Mayo scores with HBOT (**figure 3b**). We also observed a significant accompanying increase in MUC2 with HBOT in the mucosal proteomics dataset, (**figure 3c, left**) and the relative increase in MUC2 was greater in HBOT responders compared to HBOT non-responders. (**figure 3c, right**) Fecal proteomics confirmed an increase in MUC2 with HBOT, which was incremental, based on duration of exposure. (**supplementary figure 13**) In the fecal proteomic and metagenomic datasets we observed an association between reductions in *A. mucinophilia* and higher MUC protein values. (**supplementary figure 14)** HBOT response may therefore be mediated in part by the drop out of mucus digesting microbes allowing for enhanced mucus layer strength, strengthening of the epithelial barrier, and repopulation of *Firmicutes* and *Firmicute* related bile acid metabolism. Together, these effects may promote improvement in colitis.

**Figure 3:**
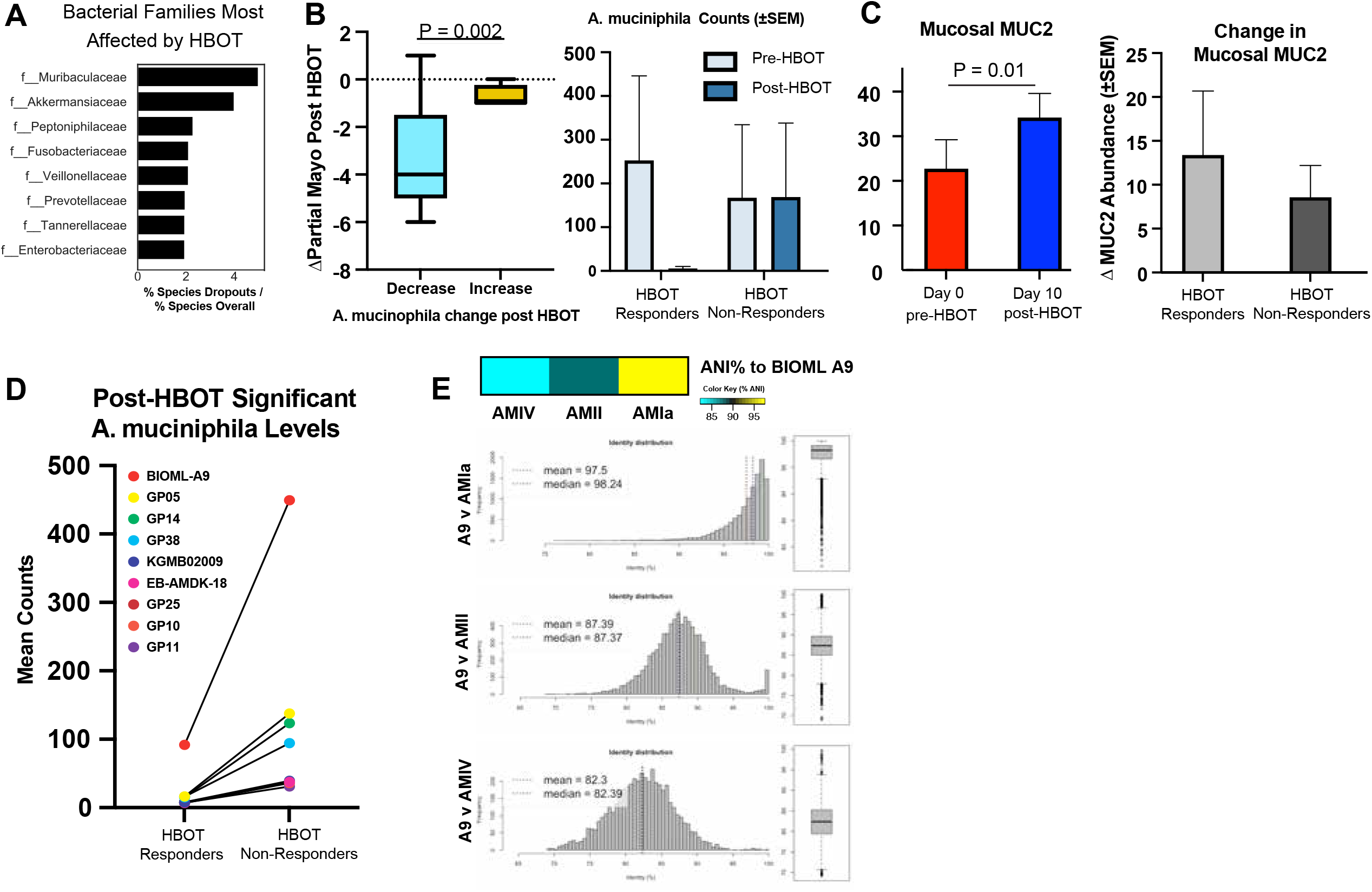
Multi-omic Characterization of Fecal Samples Identifies HBOT Response Status Associated Increases in Firmicutes and Bile Acid Production. **A:** Top 8 taxonomic families most affected by HBOT treatment calculated ratiometrically by comparing the percentage of families with species negatively altered by HBOT treatment to their overall species-level percentage representation in metagenomic features. **B:** Group-wise comparison of changes in partial Mayo score as a function of post-HBOT loss in the genus *Akkermansia* (Welch’s t-test, p = 0.014). Patients showing a decrease (< 50 counts compared to their pre-HBOT condition), or no measurable levels, were considered to be in decreased group, while no change or increase was included in the ‘increased’ group. Range represents max calculated values. **C:** Left: Group-wise comparison (paired t-test, error bars plotted using ±SEM) of mucosal proteomics abundance changes in MUC2 comparing pre-HBOT (D0, n=10) and post-HBOT (D10, n=10). Right: Group-wise comparison of post-HBOT mucosal biopsy levels of MUC2 by response group. Results were not significant (unpaired t-test p = 0.57). **D:** *A. muciniphila* strains significantly (p < 0.05, uncorrected) higher post-exposure in HBOT non-responders compared to HBOT responders. **E:** Heatmap generated using mean percentage scores of Average Nucleotide Identity (ANI) between BIOML A9 compared to several canonical phylogroups, with accompanying Histograms of ANI scores comparing BIOML A9 and several phylogroups (left and their box plot distributions (bottom).

### Strain-level variation of *A. muciniphila* comparing HBOT Non-Responders to Responders

To better characterize the lack of *A. muciniphila* reduction in HBOT non-responders, we assessed *A. muciniphila* strain variation in IBD patients. Shotgun metagenomic data was compared to 279 unique reference *A. muciniphila* strains, including strains of *A. muciniphila* previously identified to be oxygen tolerant and/or resistant (AMIa and AMII phylogroups).^22,23^ We identified 10 strains significantly greater in post-HBOT non-responders compared to the post-HBOT responders. Six of these strains (GP05, GP14, GP38, GP25, GP10, GP11) are already known to belong to the oxygen-resistant AMIa or AMII phylogroups. One of the strains not previously classified (BIOML-A9), was 5-fold higher in HBOT non-responders versus HBOT responders. (**figure 3d**) A genomic comparison of BIOML-A9 to different phylogroups demonstrated it strongly aligned with the AMIa and AMII phylogroups. (**figure 3e**) Together, these data suggest that *A. muciniphila* adaptation to oxygen exposure with ongoing mucus digestion may play an important role in treatment resistant to HBOT. **Colonization of mice with HBOT-shaped microbial communities from responders reduces colitis**

IL10^-/-^ mice demonstrate a colitis phenotype driven by NLRP3 inflammasome activity,^24^ and thus resemble phenotypes observed in UC patients. Germ-free IL10^-/-^ mice were colonized with post-treatment stool from 4 HBOT treated patients, 2 clinical responders (n=5 mice per patient sample) and 2 clinical non-responders (n=5 mice per patient sample) in order to characterize the role HBOT-resulting alterations in gut microbe composition had on host inflammation. All 4 patients had persistent endoscopic inflammation at the time of stool collection. Despite all 4 patient stool donors having active endoscopic inflammation, mice colonized with stool from the 2 HBOT clinical responders had longer colon length and less overall inflammation. (**figure 4a-c**) PCR confirmed differences in colitis activity were not related to induction of the STAT3 pathway. (**figure 4d, supplementary figure 15**) No significant differences were observed by flow cytometry analysis of T cells (data not shown). Altogether, these data demonstrate a central role played by the intestinal microbiota in driving HBOT-induced phenotypical response.

**Figure 4:**
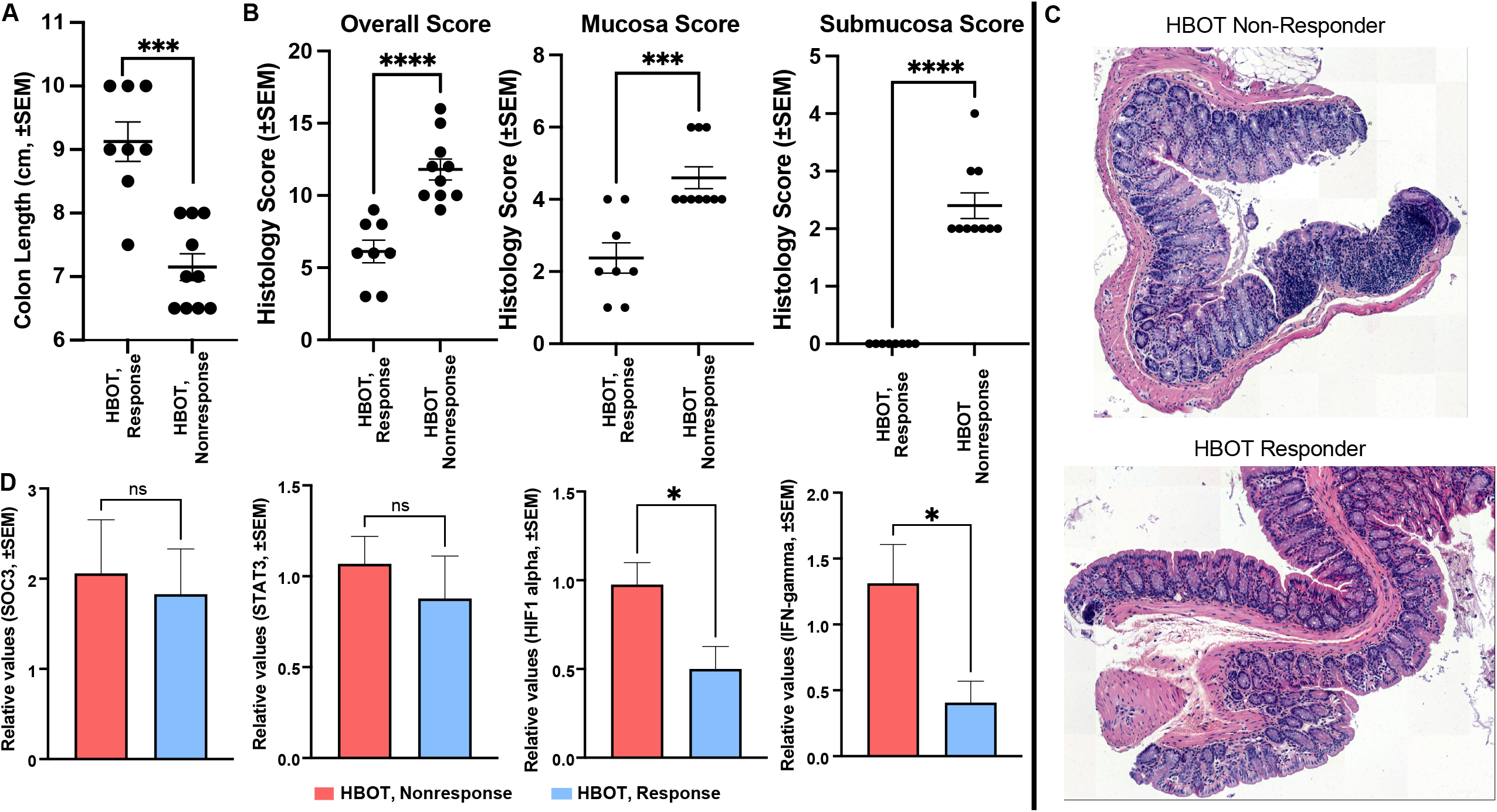
Fecal Colonization of IL10^-/-^ Mice with Stool from Responders and Non-Responders to Hyperbaric Oxygen Therapy Demonstrates Differential Modulation of Colitis. Group-wise comparison (Welch’s unpaired t-test, ±SEM) of (**A**) colon length (p=0.0004) and (**B**) histology scores between IL10^-/-^ germ-free mice colonized with post-HBOT stool from two non-responder donors (n=10 mice) and two responder donors (n=8 mice) measuring overall (e.g. summed) histological inflammation (p < 0.0001), mucosa (p=0.0004), and submucosa (p<0.0001). **C:** Representative colon histology images from mice colonized with stool from HBOT non-responders or HBOT responders. **D**: Barplot of expression differences between mouse colonized with responder or non-responder stool (n = 8 replicates, non-responders = 10 replicates) of SOCS3 (not significant), STAT3 (not significant), HIF1 (Welch’s unpaired t-test, p = 0.017), and IFN-y (Welch’s t-test, p = 0.043) as measured by qPCR on colon tissue.

## DISCUSSION

HIF-1α is important in UC pathogenesis, and targeting this may be of therapeutic benefit. Using targeted and untargeted sequencing approaches, which benefited from novel proteomics and transcriptomics technology, we identified a specific effect for HBOT on HIF-1α activity. In the epithelium HBOT resulted in significantly higher HIF-1α expression, and in neutrophil HBOT resulted in significantly lower HIF-1α expression, however, these changes were not consistent across all regions of neutrophilic infiltrate and they did not directly correlate to HBOT response status.. Furthermore, we were not able to identify consistency in associations or directionality in changes between HBOT and HIF-1α across other approaches (bulk-RNA sequencing or mucosal TMT-multiplexed proteomics). Taken together, HBOT impacts HIF-1α but changes in HIF-1α alone do not explain mechanisms underlying improvements in UC disease activity with HBOT.

Hypoxic microenvironments enhance neutrophil degranulation, particularly for azurophilic granules, and this has been thought to be partially HIF-dependent.^25^ HIF-1α is the predominant HIF sub-type found in neutrophils, however, hypoxia-augmented degranulation could not be re-demonstrated with pharmacological HIF-1α stabilization and activation alone.^25^ HIF-independent pathways involved in neutrophil degranulation under hypoxic conditions include reactive oxygen species (ROS), MAPK, and STAT3 signalling.^25,26^ Using TMT-multiplexed proteomics, we identified a consistent and highly significant association between HBOT response status and neutrophil degranulation, with specificity of effect for the STAT3-NLRP3 inflammasome related azurophilic granules in both the fecal and mucosal proteomics datasets. NLRP3 is directly linked to azurophilic granules with mutations of NLRP3 being responsible for hyper-sensitive inflammasome function disorders through an azurophilic granule-selective process.^27^ STAT3 plays a central role in NLRP3 inflammasome mediated inflammation and prior studies have demonstrated that STAT3 inhibition abrogates NLRP3 protein expression and IL-1ß release.^28,29^ In the DSP dataset, we observed a significant reduction in neutrophil STAT3 mRNA abundance, phosphorylated MAPK protein expression, and markers of ROS mediated injury. STAT3 and NLRP3 are known targets of JNK, and inhibition of MAPK signalling in neutrophils abrogates the release of azurophilic granule contents.^26,28^ At the single cell level in the proteome dataset, we observed HBOT to significantly increase the pro-apoptotic protein BIM alongside BCLXL. Hypoxia is known to prolong neutrophil survival and inhibit apoptosis,^30^ and HBOT has been shown to increase neutrophil apoptosis and enhance their clearance.^31^ BIM, a member of the Bcl-2 family, is known to promote neutrophil apoptosis and has been demonstrated to limit cytokine mediated prolonged survival of neutrophils.^32^ BCLXL mediated regulation of neutrophil apoptosis is ROS-dependent,^33^ and BCLXL has been demonstrated to inhibit NLRP3 inflammasome formation and IL-1ß activation.^34^ Prior *in-vitro* studies have demonstrated an increase in BCLXL, and reduction in ROS, MAPK, STAT3 and NLRP3, with HBOT, ^35-39^ further supporting our observations. Together these data, in combination with prior *in-vitro* studies from other groups supporting our observations, suggest that HBOT improves UC disease activity in part through the neutrophil STAT3 pathway.

Recognizing that a microbiome-neutrophil cross-talk exists,^40^ the microbiome has been observed to regulate STAT3 expression specifically,^41^ and HBOT significantly alters colonic microbial composition and function in animal models,^42^ we sought to further assess the effects of HBOT on the microbiome and whether microbial changes may have contributed to observed host effects of HBOT on STAT3. We observed an acute drop out of bacterial subpopulations post HBOT, with an accompanying proportional increase in *Firmicutes* and a key secondary bile acid, LCA. UC patients have reduced LCA, this deficiency is due to a reduction in *Firmicutes*, and LCA supplementation is protective against colitis in animal models.^43^ Several of the *Firmicutes* increased with HBOT in our trial are known producers of secondary bile acids,^44^ and in healthy volunteers hyperbaric conditions have been observed to increase bile acid producing *Firmicutes*.^45^ The specific microbes most affected by HBOT, which allowed for this proportional increase in *Firmicutes*, included known mucinophiles with an association between *Akkermansia muciniphila* and HBOT response status. *Akkermansia* is associated with risk of IBD development,^46^ but data is conflicting on whether *A. muciniphila* is pathogenic or protective against colitis.^47-49^ Strain specific anti- or pro-inflammatory properties exist for *A. muciniphila*.^50,51^ Although *A. muciniphila* is considered to be a strict anaerobe, it is capable of adapting to oxygen exposure through increased expression of respiration genes allowing it to take advantage of epithelial oxygen gradients.^22^ We observed persistence of *A. muciniphila* strains known to be oxygen tolerant and/or resistent (AMIa and AMII phylogroups) in the HBOT non-responders, suggesting adapation to oxygen may explain HBOT non-response. Furthermore, we observed an association between HBOT and MUC2 protein levels. MUC2 is the core structural protein for the inner mucus layer and weakening of this barrier is an early event in UC pathogenesis in humans and animal models (IL10^-/-^).^52^ Mutations in MUC2 determine early dysbiosis in colitis models,^53^ and MUC2^-/-^ models are characterized by increased JAK/STAT activity with specificity towards IL-6/STAT3 activity.^54,55^ MUC2 deficiency related dysbiosis is characterized by reduction in *Firmicutes* and increases in the *Muribaculaceae* and *Akkermansiaceae* families.^53^ Therefore, our observed changes with HBOT for a reduction in *Muribaculaceae* and *Akkermansiaceae* families, increase in *Firmicutes*, and increase in MUC2 are in-line with expected pathological mechanisms. Using animal models, we were able to confirm that global changes in microbial composition and metabolite alterations were associated with differential colitis induction, however, there were no differences in induction of STAT3. This suggests that HBOT response mechanisms involve direct effects on host neutrophil STAT3 pathways and separate effects on the microbiome which may further augment improvements in disease activity.

Our study has multiple strengths which include: the use of baseline and follow-up mucosal and fecal samples from two prospective clinical trials, separate healthy and disease severity matched control samples, use of novel and emerging techniques for proteomics and mRNA transcriptomics, our ability to study tissue neutrophils, and the consistency of observation and strength of associations for our proposed mechanisms of action across sequencing datasets and prior literature. Most notably we provide the first application of DSP in UC to study neutrophils and it was this approach which linked observations towards a unifying central hypothesis related to neutrophil degranulation and STAT3. Furthermore, our proposed host response mechanism is confirmed by multiple prior *in-vitro* studies by other groups and the contribution of changes in microbial composition and metabolism to improving disease activity was assessed through dedicated *in-vivo* colonization experiments. Our study is not without limitation, the most notable being the lack of in-vivo confirmation of STAT3 pathway related mechanisms for HBOT. Prior neutrophil STAT3 knockout models which developed spontaneous chronic enterocolitis also resulted in knockout of STAT3 in macrophages,^56,57^ and it was the macrophage STAT3 deficiency which contributed to development of colitis.^58^ A more neutrophilc specific STAT3 knockout model could be made through crossing STAT3^flox^ mice with MRP8-Cre mice, however, this will still result in knockout of STAT3 in up to 20% of macrophages depending on the tissue of interest.^59^ Prior work has suggested that neutrophil STAT1 activity may be a determinant of UC activity,^60^ and conditional deletion of STAT3 may result in increased activity of STAT1 through heterodimer receptor occupancy which may further compound phenotypic presentations of animal models.^61^ Further confirmation of our observations is best done within a larger phase 3 clinical trial program where comparisons of neutrophil expression profiles can be made between randomized HBOT and sham treated UC patients.

In conclusion, our multi-omics analysis has observed two complementary mechanisms through which HBOT may improve UC activity. First, we observed a reduction in neutrophil degranulation through the STAT3-NLRP3-azurophilic granule pathways. Second, we observed a decrease in mucus digesting bacteria, with an accompanying increase in MUC2, epithelial HIF-1α, *Firmicutes*, and bile acid production. Further investigation of *A. muciniphila* strain level adaptations are needed to understand mechanisms of HBOT non-response.

## Supporting information

Supplementary Results

## Data Availability

Proteomic data and supplementary files are available online at https://massive.ucsd.edu (Study ID MSV000088636). Metaproteomic data used for the severity-matched patient analyses were derived from a previous study and is available at MassIVE (Study ID MSV000083874). Metabolomics files are available at massive.ucsd.edu (Study ID: MSV000081492 and MSV000086483). Processed 16s, shotgun metagenomic and metabolomic results are available online at qiita.ucsd.edu (Study ID: 11149). Bulk RNA sequencing data is available on NCBI server (accession ID: GSE152229). All results and scripts are available at https://github.com/c6gonzalez/HBOT and https://github.com/rhmills/HBOT_Multiomics. Patient demographics are reported in Supplementary Tables 2 and 3.  

## Acknowledgements/Funding

This work was directly supported by an American Gastroenterology Association Research Scholar Award and NIDDK U34 Planning Grant (DK126626) to Parambir S. Dulai. This work was also supported by the Crohn’s and Colitis Foundation through a Litwin IBD Pioneers Program Grant, the UCSD Collaborative Center for Multiplexed Proteomics, the San Diego Digestive Disease Center (P30 DK120515), UCSD Gastroenterology T32 training grant (DK 0070202; P.S.D., M.T., R.H.M, C.S., and Y.M.), and UCSD Graduate Training Program in Cellular and Molecular Pharmacology (T32 GM007752; LR). Y.M was also supported by an NIH CTSA-funded career-development award (1TL1TR001443). PG is supported by the NIH (UG3TR003355, AI155696 and AI141630). J.T.C. is supported by the NIH (AI123202, AI129973, AI132122, and DK119724). C.G.G. is funded by Institutional Research and Academic Career Development Awards (K12GM068524). C.S. is NIH Training grant (T32, DK007202). K.F. is supported by the NIH (2UL1TR001442-06). B.C. is supported by a Starting Grant from the European Research Council (ERC) under the European Union’s Horizon 2020 research and innovation program (grant agreement No. ERC-2018-StG-804135), a Chaire d’Excellence from IdEx Université de Paris - ANR-18-IDEX-0001, an Innovator Award from the Kenneth Rainin Foundation, an ANR grant EMULBIONT ANR-21-CE15-0042-01 and the national program “Microbiote” from INSERM. P.C.D. is supported by the Crohn’s and Colitis Foundation (grant #675191).

## REFERENCES

1. Glover LE, Colgan SP. Hypoxia and metabolic factors that influence inflammatory bowel disease pathogenesis. Gastroenterology. 2011;140(6):1748–1755.

2. Colgan SP. Targeting hypoxia in inflammatory bowel disease. Journal of investigative medicine : the official publication of the American Federation for Clinical Research. 2016;64(2):364–368.

3. Dulai PS, Buckey JC, Jr., Raffals LE, et al. Hyperbaric oxygen therapy is well tolerated and effective for ulcerative colitis patients hospitalized for moderate-severe flares: a phase 2A pilot multi-center, randomized, double-blind, sham-controlled trial. Am J Gastroenterol. 2018;113(10):1516–1523.

4. Dulai PSea. A phase 2B randomized trial of hyperbaric oxygen therapy for ulcerative colitis patients hospitalized for moderate to severe flares. Alimentary pharmacology & therapeutics. 2020.

5. Helmink BA, Reddy SM, Gao J, et al. B cells and tertiary lymphoid structures promote immunotherapy response. Nature. 2020;577(7791):549–555.

6. Cabrita R, Lauss M, Sanna A, et al. Tertiary lymphoid structures improve immunotherapy and survival in melanoma. Nature. 2020;577(7791):561–565.

7. Zhang X, Deeke SA, Ning Z, et al. Metaproteomics reveals associations between microbiome and intestinal extracellular vesicle proteins in pediatric inflammatory bowel disease. Nature communications. 2018;9(1):2873–2873.

8. Campeau A, Mills RH, Blanchette M, et al. Multidimensional Proteome Profiling of Blood-Brain Barrier Perturbation by Group B Streptococcus. mSystems. 2020;5(4).

9. Elias JE, Gibbons FD, King OD, Roth FP, Gygi SP. Intensity-based protein identification by machine learning from a library of tandem mass spectra. Nat Biotechnol. 2004;22(2):214–219.

10. Kuleshov MV, Jones MR, Rouillard AD, et al. Enrichr: a comprehensive gene set enrichment analysis web server 2016 update. Nucleic Acids Res. 2016;44(W1):W90–97.

11. McDonald D, Hyde E, Debelius JW, et al. American Gut: an Open Platform for Citizen Science Microbiome Research. mSystems. 2018;3(3).

12. Thompson LR, Sanders JG, McDonald D, et al. A communal catalogue reveals Earth’s multiscale microbial diversity. Nature. 2017;551(7681):457–463.

13. Mills R, Dulai P, Vázquez-Baeza Y, et al. OP31 Meta–omics reveals microbiome-driven proteolysis as a contributing factor to the severity of ulcerative colitis disease activity. Journal of Crohn’s and Colitis. 2020;14(Supplement_1):S030–S031.

14. Ondov BD, Treangen TJ, Melsted P, et al. Mash: fast genome and metagenome distance estimation using MinHash. Genome Biol. 2016;17(1):132.

15. Zhu Q, Mai U, Pfeiffer W, et al. Phylogenomics of 10,575 genomes reveals evolutionary proximity between domains Bacteria and Archaea. Nat Commun. 2019;10(1):5477.

16. Zhu Q, Huang S, Gonzalez A, et al. OGUs enable effective, phylogeny-aware analysis of even shallow metagenome community structures. bioRxiv. 2021:2021.2004.2004.438427.

17. Parks DH, Imelfort M, Skennerton CT, Hugenholtz P, Tyson GW. CheckM: assessing the quality of microbial genomes recovered from isolates, single cells, and metagenomes. Genome Res. 2015;25(7):1043–1055.

18. Bowers RM, Kyrpides NC, Stepanauskas R, et al. Minimum information about a single amplified genome (MISAG) and a metagenome-assembled genome (MIMAG) of bacteria and archaea. Nat Biotechnol. 2017;35(8):725–731.

19. Chassaing B, Srinivasan G, Delgado MA, Young AN, Gewirtz AT, Vijay-Kumar M. Fecal lipocalin 2, a sensitive and broadly dynamic non-invasive biomarker for intestinal inflammation. PLoS One. 2012;7(9):e44328.

20. Sheshachalam A, Srivastava N, Mitchell T, Lacy P, Eitzen G. Granule protein processing and regulated secretion in neutrophils. Front Immunol. 2014;5:448.

21. Tu GW, Ju MJ, Zheng YJ, et al. CXCL16/CXCR6 is involved in LPS-induced acute lung injury via P38 signalling. J Cell Mol Med. 2019;23(8):5380–5389.

22. Ouwerkerk JP, van der Ark KCH, Davids M, et al. Adaptation of Akkermansia muciniphila to the Oxic-Anoxic Interface of the Mucus Layer. Appl Environ Microbiol. 2016;82(23):6983–6993.

23. Becken B, Davey L, Middleton DR, et al. Genotypic and Phenotypic Diversity among Human Isolates of Akkermansia muciniphila. mBio. 2021;12(3).

24. Liu L, Dong Y, Ye M, et al. The Pathogenic Role of NLRP3 Inflammasome Activation in Inflammatory Bowel Diseases of Both Mice and Humans. Journal of Crohn’s & colitis. 2017;11(6):737–750.

25. Lodge KM, Cowburn AS, Li W, Condliffe AM. The Impact of Hypoxia on Neutrophil Degranulation and Consequences for the Host. Int J Mol Sci. 2020;21(4).

26. Mócsai A, Jakus Z, Vántus T, Berton G, Lowell CA, Ligeti E. Kinase pathways in chemoattractant-induced degranulation of neutrophils: the role of p38 mitogen-activated protein kinase activated by Src family kinases. J Immunol. 2000;164(8):4321–4331.

27. Johnson JL, Ramadass M, Haimovich A, et al. Increased Neutrophil Secretion Induced by NLRP3 Mutation Links the Inflammasome to Azurophilic Granule Exocytosis. Frontiers in cellular and infection microbiology. 2017;7:507.

28. Furuya MY, Asano T, Sumichika Y, et al. Tofacitinib inhibits granulocyte-macrophage colony-stimulating factor-induced NLRP3 inflammasome activation in human neutrophils. Arthritis research & therapy. 2018;20(1):196.

29. Edwan J, Chae JJ, Goldbach-Mansky R, Colbert RA. Evidence that STAT3 controls NLRP3 inflammasome-dependent release of IL-1β and pyronecrosis through regulation of mitochondrial activity. Arthritis Rheum. 2013;65.

30. Hannah S, Mecklenburgh K, Rahman I, et al. Hypoxia prolongs neutrophil survival in vitro. FEBS Lett. 1995;372(2-3):233–237.

31. Almzaiel AJ, Billington R, Smerdon G, Moody AJ. Hyperbaric oxygen enhances neutrophil apoptosis and their clearance by monocyte-derived macrophages. Biochem Cell Biol. 2015;93(4):405–416.

32. Andina N, Conus S, Schneider EM, Fey MF, Simon HU. Induction of Bim limits cytokine-mediated prolonged survival of neutrophils. Cell Death Differ. 2009;16(9):1248–1255.

33. Perskvist N, Long M, Stendahl O, Zheng L. Mycobacterium tuberculosis promotes apoptosis in human neutrophils by activating caspase-3 and altering expression of Bax/Bcl-xL via an oxygen-dependent pathway. J Immunol. 2002;168(12):6358–6365.

34. Vince JE, De Nardo D, Gao W, et al. The Mitochondrial Apoptotic Effectors BAX/BAK Activate Caspase-3 and -7 to Trigger NLRP3 Inflammasome and Caspase-8 Driven IL-1β Activation. Cell Rep. 2018;25(9):2339-2353.e2334.

35. Vlodavsky E, Palzur E, Feinsod M, Soustiel JF. Evaluation of the apoptosis-related proteins of the BCL-2 family in the traumatic penumbra area of the rat model of cerebral contusion, treated by hyperbaric oxygen therapy: a quantitative immunohistochemical study. Acta Neuropathol. 2005;110(2):120–126.

36. Qi Y, Guo Z, Meng X, Lv Y, Pan S, Guo D. Effects of hyperbaric oxygen on NLRP3 inflammasome activation in the brain after carbon monoxide poisoning. Undersea Hyperb Med. 2020;47(4):607–619.

37. Qian H, Li Q, Shi W. Hyperbaric oxygen alleviates the activation of NLRP11311inflammasomes in traumatic brain injury. Mol Med Rep. 2017;16(4):3922–3928.

38. Selvendiran K, Kuppusamy ML, Ahmed S, et al. Oxygenation inhibits ovarian tumor growth by downregulating STAT3 and cyclin-D1 expressions. Cancer biology & therapy. 2010;10(4):386–390.

39. Grimberg-Peters D, Büren C, Windolf J, Wahlers T, Paunel-Görgülü A. Hyperbaric Oxygen Reduces Production of Reactive Oxygen Species in Neutrophils from Polytraumatized Patients Yielding in the Inhibition of p38 MAP Kinase and Downstream Pathways. PLoS One. 2016;11(8):e0161343.

40. Zhang D, Frenette PS. Cross talk between neutrophils and the microbiota. Blood. 2019;133(20):2168–2177.

41. Li Y, Kundu P, Seow SW, et al. Gut microbiota accelerate tumor growth via c-jun and STAT3 phosphorylation in APCMin/+ mice. Carcinogenesis. 2012;33(6):1231–1238.

42. Albenberg L, Esipova TV, Judge CP, et al. Correlation between intraluminal oxygen gradient and radial partitioning of intestinal microbiota. Gastroenterology. 2014;147(5):1055-1063.e1058.

43. Sinha SR, Haileselassie Y, Nguyen LP, et al. Dysbiosis-Induced Secondary Bile Acid Deficiency Promotes Intestinal Inflammation. Cell Host Microbe. 2020;27(4):659-670.e655.

44. Guo P, Zhang K, Ma X, He P. Clostridium species as probiotics: potentials and challenges. Journal of animal science and biotechnology. 2020;11:24.

45. Oya M, Tadano Y, Takihata Y, et al. Effects of hyperbaric conditions on fecal microbiota. Bioscience of microbiota, food and health. 2019;38(1):35–39.

46. Zhang ZJ, Qu HL, Zhao N, et al. Assessment of Causal Direction Between Gut Microbiota and Inflammatory Bowel Disease: A Mendelian Randomization Analysis. Front Genet. 2021;12:631061.

47. Earley H, Lennon G, Balfe Á, Coffey JC, Winter DC, O’Connell PR. The abundance of Akkermansia muciniphila and its relationship with sulphated colonic mucins in health and ulcerative colitis. Scientific Reports. 2019;9(1):15683.

48. Bian X, Wu W, Yang L, et al. Administration of Akkermansia muciniphila Ameliorates Dextran Sulfate Sodium-Induced Ulcerative Colitis in Mice. Frontiers in Microbiology. 2019;10(2259).

49. Seregin SS, Golovchenko N, Schaf B, et al. NLRP6 Protects Il10(-/-) Mice from Colitis by Limiting Colonization of Akkermansia muciniphila. Cell Rep. 2017;19(10):2174.

50. Zhai R, Xue X, Zhang L, Yang X, Zhao L, Zhang C. Strain-Specific Anti-inflammatory Properties of Two Akkermansia muciniphila Strains on Chronic Colitis in Mice. Frontiers in cellular and infection microbiology. 2019;9(239).

51. Liu Q, Lu W, Tian F, et al. Akkermansia muciniphila Exerts Strain-Specific Effects on DSS-Induced Ulcerative Colitis in Mice. Frontiers in cellular and infection microbiology. 2021;11(710).

52. van der Post S, Jabbar KS, Birchenough G, et al. Structural weakening of the colonic mucus barrier is an early event in ulcerative colitis pathogenesis. Gut. 2019;68(12):2142–2151.

53. Liso M, De Santis S, Verna G, et al. A Specific Mutation in Muc2 Determines Early Dysbiosis in Colitis-Prone Winnie Mice. Inflammatory bowel diseases. 2020;26(4):546–556.

54. Lu P, Burger-van Paassen N, van der Sluis M, et al. Colonic gene expression patterns of mucin Muc2 knockout mice reveal various phases in colitis development. Inflammatory bowel diseases. 2011;17(10):2047–2057.

55. Shan YS, Hsu HP, Lai MD, et al. Suppression of mucin 2 promotes interleukin-6 secretion and tumor growth in an orthotopic immune-competent colon cancer animal model. Oncol Rep. 2014;32(6):2335–2342.

56. Takeda K, Clausen BE, Kaisho T, et al. Enhanced Th1 activity and development of chronic enterocolitis in mice devoid of Stat3 in macrophages and neutrophils. Immunity. 1999;10(1):39–49.

57. Kasembeli MM, Bharadwaj U, Robinson P, Tweardy DJ. Contribution of STAT3 to Inflammatory and Fibrotic Diseases and Prospects for its Targeting for Treatment. International journal of molecular sciences. 2018;19(8).

58. Reindl W, Weiss S, Lehr HA, Förster I. Essential crosstalk between myeloid and lymphoid cells for development of chronic colitis in myeloid-specific signal transducer and activator of transcription 3-deficient mice. Immunology. 2007;120(1):19–27.

59. Stackowicz J, Jönsson F, Reber LL. Mouse Models and Tools for the in vivo Study of Neutrophils. Front Immunol. 2019;10:3130.

60. Schreiber S, Rosenstiel P, Hampe J, et al. Activation of signal transducer and activator of transcription (STAT) 1 in human chronic inflammatory bowel disease. Gut. 2002;51(3):379–385.

61. Qing Y, Stark GR. Alternative activation of STAT1 and STAT3 in response to interferon-gamma. J Biol Chem. 2004;279(40):41679–41685.

