## Supplementary Methods for "Ulcerative Colitis Host-Microbiome Response to Hyperbaric Oxygen Therapy"

### Immunohistochemistry

IHC was performed for HIF-1α and HO-1 with an *apriori* hypothesis that HBOT response status would correlate to changes in these pathways, thereby demonstrating these to be central to HBOT mechanisms in UC. Post-hoc IHC was done for STAT3 and phosphorylated STAT3 to confirm observations made through digital spatial profiling and proteomics.

FFPE tissue sections of 4 µm thickness were cut and placed on glass slides coated with poly-L-lysine, followed by deparaffinization and hydration. Heat-induced epitope retrieval was performed using citrate buffer (pH 6.0) in a pressure cooker. Tissue sections were incubated with 0.3% hydrogen peroxidase for 15 m to block endogenous peroxidase activity, followed by incubation with primary antibodies for overnight in a humidified chamber at 4°C. Antibodies used for immunostaining; anti- HIF-1α [EP1215Y] [1:100, Rabbit monoclonal antibody], anti- Anti-Heme Oxygenase 1 antibody [EP1391Y] [1:50, rabbit monoclonal antibody]. Immunostaining was visualized with a labeled streptavidin-biotin using 3,3′-diaminobenzidine as a chromogen and counterstained with hematoxylin. Samples were quantitatively analyzed and scored based on presence (positive) or absence (negative) of staining by a blinded pathologist with expertise in gastrointestinal diseases. Data is displayed as frequency of staining score and a Chi-square test was used to determine significance.

Immunohistochemical slides were cut at 4um from formalin-fixed paraffin-embedded tissue and air dried at room temp before baking at 60 degrees Celsius for 30 minutes. Automated protocol performed on the Leica Bond Rx and includes paraffin dewax, antigen retrieval and staining. Heat induced epitope retrieval using Bond Epitope Retrieval 1, pH6 (Leica Biosystems AR9640) or Bond Epitope Retrieval 1, pH9 (Leica Biosystems AR9961) was incubated at 100 degrees Celsius for 20 minutes. Primary antibody staining was applied and incubated at room temp for 30 minutes for STAT 3 (1:800). Primary antibody binding is detected and visualized using the Leica Bond Polymer Refine Detection Kit (DS9800) with DAB chromogen and Hematoxylin counterstain. The automated Stainer/Detection system used was the Leica Biosystems Inc. and the antibodies used included: Phospho-Stat3 (Tyr705, D3A7, XP® Rabbit mAb #9145, 1:400 dilution), and Stat3 (D3Z2G, Rabbit mAb #12640, 1:800 dilution).

### Bulk RNA Sequencing

Bulk-RNA sequencing was done for pre- and post-intervention mucosal biopsies from the sham-controlled phase 2A trial. Biopsies were immediately frozen and stored at -80 degrees Celsius. Biopsy samples were homogenized manually on ice in TRIzol Reagent (Thermo Fisher Scientific) and processed according to the manufacturer's instructions. cDNA libraries were prepared using TruSeq non-Stranded Total RNA Sample Prep Kit (Illumina) according to manufacturer's instructions. cDNA libraries were sequenced with a HiSeq2500 (Illumina) at the University of California San Diego Institute for Genomic Medicine facility.

RNAseq reads were processed by first removing adapter sequences with cutadapt (1.14.0) and mapped to a database of repetitive elements (RepBase 18.05) using STAR (2.4.0i). Reads that went unmapped to repeat elements were then mapped to the human genome (hg19) using default parameters. Aligned reads were annotated using featureCounts (v1.5.3) with Gencode V19 annotations. Differential expression was performed with DESeq2 (1.22.1). GO enrichment was performed against significant (p-adj <= 0.05) dysregulated genes using a background of all expressed genes (average DESeq2 normalized count >= 1). Ontology data was downloaded from the Ensembl (GRCh37) Biomart API.

**Digital Spatial Profiling**

The GeoMx Digital Spatial Profiler (DSP) developed by NanoString Technologies enables spatially resolved, high-plex (10s -10,000s) digital quantitation of proteins and mRNA in tissue. The assay utilizes unique reagents (antibodies or RNA probes) coupled to UV photocleavable oligonucleotide barcodes. After incubation and hybridization of the GeoMx reagents along with visualization reagents to slide-mounted FFPE tissue, the DSP scans the slides and presents a high-quality image for region of interest selection. The oligonucleotide tags are released from user-selected regions of the tissue with focused UV light based on the user region and segmentation selections. The digital micromirror device of the DSP instrument tunes the UV light with 1□m resolution, allowing great flexibility for region selection including irregularly shaped and non- continuous segments. Released tags are quantitated with nCounter optical barcodes or an Illumina next- generation sequencer. Counts are mapped back to the tissue location by the software, resulting in a spatially- resolved digital profile of protein or mRNA abundance. For the current study, protocols were followed according to the manufacturer's instructions and in accordance with prior published work using this platform.[5,6] Briefly, a 5-10μm thick FFPE tissue section was stained with oligo-tagged antibodies to detect a predetermined panel of markers. (**Supplementary Table 1**) Oligo tags are attached to the antibody and ISH probes via a UV- photocleavable linker. Regions of interest (ROI) were selected on the visualized tissue based on cell positivity for elastase and CD45. After ROI selection, oligos from the selected regions were released upon focused exposure to UV light using the Nanostring DSP instrument and photocleaved oligos are aspirated via a microcapillary tube and stored in microplate wells. Lastly, the photocleaved oligos from the spatially-resolved ROI in the microplate were quantified using next-generation sequencing Illumina workflows.

Data were pre-processed in Nanostring DSP Software v 2.1.0.106 normalizing to ERCC counts and housekeeping genes to adjust for systemic variations to enable expression counts to be compared across genes and samples. Quality control metrics were assessed and two low quality regions were removed due to low nuclei counts < 20. The distribution of the pre-processed count data reflected a negative binomial distribution as expected, with a need for further normalization with TMM and voom. After TMM and voom normalization, the data met the assumptions necessary for downstream statistical analyses including differential expression analysis using limma-voom accounting for within case repeated measures (ROIs) and spatial deconvolution analysis.

The R BioConductor packages edgeR and limma were used to implement the limma-voom method for differential expression analysis. We normalized and analyzed Neutrophil and Immune Cell ROIs separately and subset RNA only and Protein only for downstream analysis. Trimmed mean of M-values (TMM) normalization was applied. The experimental design was modeled upon treatment and time accounting for repeated measures using consensus correlation with the design (~0 + treatment_group_time). The voom method was employed to model the mean-variance relationship, after which lmFit was used to fit per-gene linear models and empirical Bayes moderation was applied with the eBayes function. Significance was defined by using an adjusted p-value cut-off of <0.05 after multiple testing correction using a moderated t-statistic in limma. Cell type deconvolution was performed using the log-normal regression algorithm of Danaher et al. and its implementation in Bioconductor. Inputs were the normalized expression values (cpm), the background matrix of background values set to a value of 6, and the training matrix containing log2 expression values in the training set, which is part of the R package and named “Human_Cell_Landscape”.

**Proteomics**

*Mucosal Proteomics*

Each biopsy specimen was initially isolated from storage buffer and 500 uL of lysis buffer was added (7% SDS, 50 mM Tris, 6M urea, pH 8.1). Samples were then sonicated for 10 s. on 10 s. off cycles for approximately six cycles or until specimen was completely lysed (QSonica). Samples were then reduced with 10 uL of 50 mM dithiothreitol (Sigma-Aldrich) for 30 minutes at 56ºC and subsequently alkylated with 30 uL of 50 mM Iodoacetamide (Sigma-Aldrich) for 1 hour at room temperature in the dark. To each sample, S-trap “binding buffer” (PBB, a 7:1 solution of 90% methanol, 10% 1M TEAB pH adjusted to 7.1 with phosphoric acid) was added along with 50 uL 12% phosphoric acid. Samples were washed 5x with PBB and digested with 5 ug trypsin (for 3 hrs. at 47 ºC, as described in the S-trap protocol. https://cdn.shopify.com/s/files/1/0271/1964/8832/files/S-Trap_micro_protocol_long.4.6.pdf. Peptides were then eluted with 50 uL 50 mM TEAB, followed by 50 uL of 0.2% formic acid (FA), and finally 100 uL of 50% acetonitrile, 0.2% FA. Eluate was captured and the volume was evaporated using a Thermo Scientific Speedvac. Dried samples were then resuspended 500 uL 0.2% FA and desalted using Seppak tC18 cartridges and subsequently dried down (Waters WAT036820). Each sample was then resuspended in 1000 uL 50% acetonitrile (ACN) and peptide concentration was determined using Pierce™ Quantitative Colorimetric Peptide Assay (Catalog # 23275). 50 ug of peptide was dried down and labeled using TMT10plex™ Isobaric Labeling reagents (Catalog # 90406) as described in Campeau et al.[7] Labeled peptides were subsequently dried down, desalted, and fractionated using high pH reverse phase fractionation (Thermo Ultimate 3000 HPLC off-line fraction collector and c18 column, 37 s. per well, 96 wells total). After fractionation samples were concatenated into 24 samples. 12 fractions were used in the analysis of each sample.

Labeled peptides were resuspended in 8 uL 5% FA, 5% ACN wherein 3 uL was loaded onto an in- house laser-pulled 100 um ID nanospray column packed to ~30 cm with 1.8um C18 beads (Sepax technologies). Peptides were separated by reversed-phase chromatography on a Thermo Easy Nano-LC for 3 hr. per fraction. Buffer A of the mobile phase contained 0.1% formic acid (FA) in HPLC-grade water, while buffer B contained 0.1% FA in acetonitrile (ACN). The HPLC flow rate was 0.300 uL/minute. Samples were run on either a Thermo Fusion mass spectrometers that collected MS data in positive ion mode within the 600- 1200 m/z range (+2 charge), or 500-1200 m/z (+3/4 charge). A top-10 MS3 method was employed on the Fusion with an initial Orbitrap scan resolution of 60,000. This was followed by high-energy collision-induced dissociation in the ion trap and subsequent reporter ion analysis in the orbitrap. Dynamic exclusion was enabled (repeat count of 1, exclusion duration of 90 s.). The automatic gain control for FT full MS was set to 4e5 and for ITMSn was set to 1e4. ITCID was used at $MS^2$ method and the $MS^3$ AGC was set to 2e5.

Spectra were searched using Proteomics Discoverer 2.1 and the SEQUEST algorithm using a target decoy strategy as previously described[8]. Static modifications were set to carbamidomethylation (Δ57.021) and TMT tags (Δ229.163), while dynamic modifications were set to methionine oxidation (Δ15.995) and phosphorylation (Δ79.966) of serine, threonine, or tyrosine. Precursor tolerance was set to 50.00 ppm while fragment tolerance was set to 0.6 Da. Resulting peptide spectral matches were filtered at a 0.01 FDR by the Percolator module against the decoy database. Peptide intensities were summed to the protein level.

Statistics were calculated and visualized using R and the following packages: ggplot2 2.2.1, Hmisc 4.0- 3, psych 1.7.8, ggpubr 0.1.5, RColorBrewer 1.1-2. Univariate statistical analysis ($p < 0.05$) for HBOT (repeated measures t-test) samples and the public dataset (independent t-test) was done by GraphPad Prism 9.0.0. Protein abundance was normalized to a pooled TMT “bridge” channel. Additionally, each channel was further median scaled. Proteins with more than 50% of abundance values initially present in pre- and post-HBOT samples were subjected to imputation using the missForest R package (v1.4). Enrichment analysis was done by first recording statistically significant ($p < 0.05$, paired significance-test) proteins that were negatively correlated with disease severity score post-HBOT, and submitting this list of proteins to Enrichr, where the calculated “Combined Enrichment Score” was visualized[9].

*Fecal Proteomics*

Fecal samples were measured out to ~0.5 g and suspended in 5 mL of ice-cold, sterile TBS. Samples were vortexed until completely suspended. Two 20 µM vacuum, steriflip (Milipore) filters were used per sample to remove particulate. Cells were pelleted through centrifugation at 4000 rpm for 10 min at 4 ºC. Next, cells were lysed in 2 mL of buffer containing 75 mM NaCl (Sigma), 3% sodium dodecyl sulfate (SDS, Fisher), 1 mM NaF (Sigma), 1 mM beta-glycerophosphate (Sigma), 1 mM sodium orhtovanadate (Sigma), 10 mM sodium pyrophosphate (Sigma), 1 mM phenylmethylsulfonyl fluoride (PMSF, Sigma), and 1X Complete Mini EDTA-free protease inhibitors (Roche) in 50 mM HEPES (Sigma), pH 8.5. An equal volume of 8M Urea in 50 mM HEPES, pH 8.5 was added to each sample. Cell lysis was achieved through two 15-second intervals of probe sonication at 25% amplitude. Proteins were then reduced with dithiothreitol (DTT, Sigma), alkylated through iodoacetamide (Sigma), and quenched. Proteins were next precipitated via chloroform-methanol precipitation and protein pellets were dried. Protein pellets were re-suspended in 1M urea in 50 mM HEPES, pH 8.5 and digested overnight at room temperature with LysC (Wako). A second, 6-hour digestion using trypsin at 37 ºC was performed and the reaction was stopped through addition of 10% trifluoroacetic acid (TFA, Pierce).

Samples were then desalted through C18 Sep-Paks (Waters) and eluted with a 40% and 80% Acetonitrile solution containing 0.5% Acetic Acid. Concentration of desalted peptides was determined, and 50 µg aliquots of each sample were dried in a speed-vac. Additionally bridge channels consisting of 25 µg from each sample were created and 50 µg aliquots of this solution were used in the 126 channel for each Tandem Mass Tag (TMT, Thermo Fisher Scientific) 10 plex MS experiment. These bridge channels were used to control for labeling efficiency, inter-run variation, mixing errors and the heterogeneity present in each sample. Mass defects for each TMT set were accounted for in the database searches according to manufacturer’s report per lot number. The lot numbers for TMT reagents were TA262347 for the first cohort of samples and VA296083 for the second cohort of samples. Each sample or bridge channel was resuspended in 30% dry acetonitrile in 200 mM HEPES, pH 8.5 for TMT labeling with 8 µL of the appropriate TMT reagent. Reagents in the 126 channels were used to bridge between mass spec runs while remaining reagents were used to label samples in random order. Labeling was carried out for 1 hour at room temperature, and quenched by adding 9 µL of 5% hydroxylamine (Sigma). Labeled samples were acidified by adding 50 µL of 1% TFA. After TMT labeling each 10-plex experiment was combined and desalted through C18 Sep-Paks and dried in a speed-vac.

Basic pH reverse-phase liquid chromatography (LC) followed by data acquisition through LC-MS$^2$/MS$^3$ was performed. Briefly, 60-minute linear gradients of acetonitrile were performed on C18 columns using an

Ultimate 3000 HPLC (Thermo Scientific). Subsequently, 96 fractions were combined, and further separation of fractions was performed with an in-line Easy-nLC 1000 (Thermo Fisher Scientific) and a chilled autosampler. LC-$MS^2$/$MS^3$ data was collected on an Orbitrap Fusion (Thermo Fisher Scientific) mass spectrometer with acquisition and separation settings.

Data was processed using Proteome Discoverer 2.1 (Thermo Fisher Scientific). $MS^2$ data was searched against both a public repository of microbial gut genes and the human proteome (www.uniprot.org). The Sequest searching algorithm was used to align spectra to database peptides. A precursor mass tolerance of 50 parts per million (ppm) was specified and 0.6 Da tolerance for $MS^2$ fragments. Included in the search parameters was static modification of TMT 10-plex tags on lysine and peptide n-termini (+229.162932 Da), carbamidomethylation of cysteines (+57.02146 Da), and variable oxidation of methionine (+15.99492 Da). Raw data was searched at a peptide and protein false discovery rate of 1% using a reverse database search strategy. A second search was performed taking proteins assigned to either the forward or reverse database into a subset database in order to increase the spectral match rate within complex data types like the fecal proteome.

TMT reporter ion intensities were extracted from $MS^3$ spectra for quantitative analysis and signal-to- noise values were used for quantitation. Additional stringent filtering was used removing any moderate confidence peptide spectral matches (PSMs), or ambiguous PSM assignments. Additionally, any peptides with a spectral interference above 25% were removed, as well as any peptides with an average signal to noise ratio less than 10. All signal from PSMs assigned to the same protein group were summed to represent protein abundance.

Protein relative abundances were normalized first to the pooled standards for each protein and then to the median signal across the pooled standard. An average of these normalizations was used for the next step. To account for slight differences in amounts of protein labeled, these values were then normalized to the median of the entire dataset and reported as final normalized summed signal-to-noise ratios per protein per sample. Hits to microbial proteins were removed to focus analyses toward host proteins. Proteomic datasets generated from IBD patient samples resulted in final data tables containing 1,803 proteins for cohort 1 fecal samples and 2,928 proteins for cohort 2 fecal samples.

Data analysis was performed in python, and records of the code are available in corresponding Jupyter Notebooks for this project (https://github.com/rhmills/HBOT_Multiomics). Individual protein abundances from day 1 samples were compared to the abundances of day 10 samples using independent t-tests of unequal variance in scipy (www.scipy.org). Enriched or depleted proteins were determined by π–score, which accounts for both fold change and p-value. A statistical cutoff for highly ranked associations was set to $|\pi| > 1$, as previously performed. Datasets generated for the two cohorts of patients were analyzed independently. Human protein gene functional enrichment analysis was performed using DAVID, with all human proteins identified as a background list. The python package, Seaborn (version 0.9.0) was used to generate bar plots comparing the -log10 transformed FDR p-values provided by DAVID for each functional grouping. To account for protein changes related to severity, data was analyzed from a prior project of our group with data available online at www.massive.ucsd.edu (study ID MSV000082094). For this, an equal number of UC patient samples were analyzed by identical methods using a subset of samples with the same average disease activity (determined by partial Mayo scores) as the day 1 and day 10 samples from the hyperbaric oxygen- treated patients of this study. A final enrichment score was compiled for each term by comparing the –log10 transformed FDR p-values related

HBOT treatment and subtracting the –log10 transformed FDR p-values related to the mock severity-matched samples.

Protein-protein interaction networks were created through STRING-db. The association of each protein to hyperbaric oxygen treatment was determined by subtracting the π of proteins in the mock severity-matched samples to the π related to hyperbaric-oxygen treatment for proteins identified in both groups. These composite scores were determined as increased or decreased more than expected by using a cutoff of |1|. All significant associations were analyzed for protein-protein interactions using networks created through STRING-db. Associations between proteins were determined through default settings, accounting for textmining, experiments, databases, co-expression, neighborhood, gene fusion and co-occurrence. After identifying a strong association to neutrophil degranulation, proteins specific to this subnetwork were further analyzed. Connections were restricted to interactions between proteins within the query list only. Networks
were subsequently visualized through Cytoscape (version 3.5.1). Edges within protein networks were based on the combined evidence scores, with the transparency of the edge indicating higher confidence in the association. Granule type associations were determined by comparing the percentage of significant or not significant proteins that each granule type occupied in relation to the percentage of proteins that each granule type occupied among the total list of human proteins identified (% Expected). Ratios were log2 transformed and plotted through a Seaborn bar plot.

**16S gene amplicon sequencing**

Frozen samples were thawed and transferred into 96-well plates containing garnet beads and extracted using Qiagen MagAttract DNA kit adapted for magnetic bead purification. DNA was eluted in 100 µl Qiagen elution buffer. 16S rRNA gene amplicon sequencing was performed according to the Earth Microbiome Project. Briefly, the V4 region of the 16S rRNA gene (515f/806r) was amplified from 1 ul DNA per sample in triplicate. Amplicons were quantified with Quant-iT™ PicoGreen™ dsDNA Assay Kit, and 240 ng, or maximum 15 ul, of each sample was pooled into a final library and cleaned using the QIAquick PCR Purification Kit. Paired-end sequencing was performed on the Illumina MiSeq using MiSeq Reagent Kit v3 (300-cycle). 16S fastq were split, demultiplexed, trimmed to 150 base pairs, and processed through deblur to generate amplicon sequencing varients (ASVs) using QIITA (Study ID 11149).

**Metabolomics**

Frozen human stool samples were lyophilized using a CentriVap Benchtop Centrifugal Vacuum Concentrator (Labconco) attached to a Savant Ultra Low Pressure Refrigerated Vapor Trap RVT5105 (Thermo Scientific). Dried stool samples were weighed out to 30 mg (± 0.5 mg) dried weight and 1 mL extraction solvent (1:1 methanol:water spiked with 0.625 µM sulfamethazine) was added to each sample. Fecal samples were homogenized for 5 min at 25 Hz using a TissueLyser II (Qiagen), followed by a 15 minute centrifugation at 14,000 rpm. 900 µL of supernatant were transferred to new microcentrifuge tubes, which where snap frozen at -80 °C prior to another lyophilization. Fecal metabolite extracts were resuspended with 150 µL of resuspension solvent (1:19 acetonitrile:water spiked with 1 µM sulfadimethoxine), transferred to sample injection vials, and stored at 4°C prior to LC-MS/MS analysis.

Untargeted metabolomics analysis was performed using an ultra high performance liquid chromatography system (Vanquish, Thermo Scientific) coupled to a quadrupole-Orbitrap mass spectrometer (Q Exactive, Thermo

Scientific). A Phenomenex Kinetex column ( C18 1.7 µm, 2.1 mm x 50 mm) and a mobile flow-rate of 0.500 mL was used for all analysis. A 5 µL injection volume was used for all samples. The mobile phase composition was: (A) 100% LC-MS grade water spiked with 0.1 % formic acid (v/v), (B) 100% LC-MS grade acetonitrile spiked 0.1 %formic acid (v/v). The chromatographic gradient was: 0.0–1.0 min, 5% B; 1.0– 9.0 min, 5–100% B; 9.0-11.0 min, 100% B; 11.0-11.5 min, 100-5% B; 11.5-12.5 min, 5% B. The following heated electrospray ionization parameters were used: auxiliary gas flow rate, 14.0 (arb. units); auxiliary gas heater temperature, 435.0 °C; capillary temperature, 268.0 °C; sheath gas flow rate, 52.0 (arb. units); spray voltage, 3.5 kV; and S-lens RF, 50 (arb. units). Positive mode MS data was acquired using a data dependent acquisition method where the five most abundant ions are identified and subsequently scanned for MS/MS fragmentation via collision-induced dissociation. MS1 data was collected at a resolution of 35,000 and spanned an m/z range of 100-1500. MS2 data was collected at a resolution of 17,500.

Thermo proprietary ms files(.raw) were converted to a GNPS compatible format (.mzXML) using the Proteowizard program MSConvert. The open-source software MZmine version 2.37 was used for feature detection. MZmine modules were used with the following settings. Mass detection (1E5 MS1 noise level, 1E2 MS2 noise level, MS1 min time span (min) = 0.05, MS1 min height = 3E5, MS1 m/z absolute tolerance = 0.005, MS1 ppm tolerance = 10 ppm), deconvolution (min peak height = 5E3, Peak duration = 0-10 min, m/z range for MS2 scan pairing = 0.005 Da, RT range for MS2 scan pairing = 0.2 min), isotope grouper (m/z absolute tolerance = 0.05 Da or 10 ppm, RT tolerance = 0.1 min, Maximum charge = 4, representative isotope = Most intense), join aligner (m/z tolerance = 0.005 Da or 10 ppm), rows filter (Minimum peaks in a row = 2), gap filling (intensity tolerance = 0.2, m/z tolerance = 0.005 Da or 10 ppm, RT tolerance = 0.1 min).

**Metagenomics**

Metagenomic samples were initially extracted as dictated by the Earth Microbiome Project protocol using a Qiagen MagAttract PowerSoil DNA kit as previously described. Briefly, swabbed fecal material was plated into 96-well PowerBead DNA plates containing garnet beads. DNA was extracted in accordance with manufacturer's suggested protocol, with an additional incubation at 65°C for 10 min following the addition of lysis solution and immediately prior to shaking (Qiagen TissueLyser II – product 85300). The KingFisher Flex automated system was used for magnet-based DNA purification of all samples. From the purified samples, whole-genome shotgun libraries were generated using Kapa Hyperplus DNA library preparation kits (Roche, Indianapolis, IN, USA) and a 1:10 miniaturized-reaction volume. Libraries were subsequently sequenced using Illumina NovaSeq paired-end sequencing and processed using the Qiita processing and analysis platform.

**_Akkermansia_ spp. targeted database**

A set of *Akkermansia* spp. genomes were selected from NCBI RefSeq and GenBank databases on October 8th, 2021. In order to assess the completeness and contamination of the Akkermansia genomes, we used the lineage-specific workflow from CheckM v1.1.02 with the default database. We then performed a quality filtering of genomes (contamination > 5% and completeness < 90%). Closely-related genomes may present a problem for our database because they increase the ambiguity during read classification. Therefore, we identified and removed such genomes. For this, we first estimated the distance between high-quality genomes with Mash v1.13, testing multiple k-mer sizes. We then performed a hierarchical clustering over the mash distance matrix using the average linkage method, implemented in SciPy v1.7.1. Considering the strains of interest for this study, we chose an optimal distance cut-off value to determine the number of clusters. We then selected a representative genome for each cluster as the genome with the lowest distance to other genomes of the same cluster. Finally, we merged the high-quality and non-redundant *Akkermansia* spp. genomes with the current Web of Life (WoL) database.

**Sequence alignment and classification of _Akkermansia_ strains**

FASTQ sequences of studies 11149 and 12675 were downloaded from the Qiita web server. We aligned all sequences against the updated WoL database, containing the high-quality and non-redundant *Akkermansia* spp. genomes with Bowtie 2 v2.4.4. Woltka v0.1.3 was used to classify aligned sequences at the species and strain level, filtering features (species and strains) whose abundance in particular samples was below 0.01% since very low-abundance hits are false-positive assignments.

**Fecal microbiota transplantation**

Germ-free C57BL/6 IL10$^{-/-}$ male mice (C57BL/6NTac-*Il10*$^{em8Tac}$; Taconic reference GF-16006) were maintained in isolated ventilated cages Isocages (Techniplast, West Chester, PA, USA). At 6-7 weeks of age, mice were orally administered with 200 µL of fecal suspension from 2 patients who responded clinically to HBOT and 2 patients who did not respond to HBOT. Transplanted mice were group-housed (n=5) in isolated ventilated cages and fed autoclaved Purina Rodent Chow # 5021 at Cochin Institute (INSERM U1016 Paris, France) under institutionally approved protocols (APAFIS#24788-2019102806256593 v8). Mice were weighed and fresh feces were collected at weeks four and eight. Eight weeks post microbiota transplantation, mice were weighed, euthanized, and tissue and feces were collected for further analysis.

<u>H&E staining of colonic tissue and histopathologic analysis</u>

Mouse colons were fixed in 4% PFA solution and embedded in paraffin. Tissues were sectioned at 5-µm thickness and stained with hematoxylin & eosin (H&E) using standard protocols. Images were acquired using a Lamina (Perkin Elmer) at the Hist'IM platform (INSERM U1016, Paris, France). Histological scoring was determined on each colon as previously described. Briefly, each colon was assigned four scores based on the degree of epithelial damage and inflammatory infiltrate in the mucosa, submucosa and muscularis/serosa. Each of the four scores was multiplied by a coefficient 1 if the change was focal, 2 if it was patchy and 3 if it was diffuse and the 4 individual scores per colon were added.

<u>Quantification of fecal lipocalin-2 (Lcn-2) by ELISA</u>

For quantification of fecal Lcn-2 by ELISA, frozen fecal samples were reconstituted in PBS containing to a final concentration of 100 mg/mL and vortexed for 20 min to get a homogenous fecal suspension. These samples were then centrifuged for 10 min at 14,000 g and 4°C. Clear supernatants were collected and stored at -20°C until analysis. Lcn-2 levels were estimated in the supernatants using Duoset murine Lcn-2 ELISA kit (R&D Systems, Minneapolis, MN, USA) using the colorimetric peroxidase substrate tetramethylbenzidine, and optical density (OD) was read at 450 nm (SpectraMax ABS Plus microplate reader, Molecular Device).

Colonic RNAs extraction and q-RT-PCR analysis

Distal colon was collected during euthanasia and placed in RNAlater. Total RNAs were isolated from colonic tissues using TRIzol (Invitrogen, Carlsbad, CA) according to the manufacturer's instructions and as previously described. Quantitative RT-PCR were performed using the QIAGEN kit QuantiFast SYBR Green RT-PCR in a CFX96 apparatus (Bio-Rad, Hercules, CA) with specific mouse oligonucleotides (Table below). Gene expressions are presented as relative values using the DDCt approach with 36B4 housekeeping gene as reference.

| Gene | Forward primer | Reverse primer |
|---|---|---|
| ***Shp*** | AGGAACCTGCCGTCCTTCTG | CTCAGCCACCTCGAAGGTCA |
| ***Fxr*** | CCTGAGAACCCACAGCATTT | GTGTCCATCACTGCACATCC |
| ***Socs3*** | GGAACTTGTTTGCGCTTTGATT | TCACACACCCTTTTCTCTTCCAT |
| ***Stat3*** | CTTGTCTACCTCTACCCCGACAT | GATCCATGTCAAACGTGAGCG |
| ***Hif1*** | ATCAAGTCAGCAACGTGGAA | AATGGGTTCACAAATCAGCAC |
| ***TNF*** | AGGCTGCCCCGACTACGT | GACTTTCTCCTGGTATGAGATAGCAAA |
| ***IL-6*** | ACAAGTCGGAGGCTTAATTACACAT | TTGCCATTGCACAACTCTTTTC |
| ***IL-17*** | TGAGCTTCCCAGATCACAGA | TCCAGAAGGCCCTCAGACTA |
| ***IFN*** | AGCTGCAGGCCTTCAAAAAG | TGGGAGTGAATGTGGCTCAG |

Cells isolation and Flow Cytometry

Spleen and mesenteric lymph nodes (MLN) cells were collected after euthanasia in HBSS. Tissues were then grinded successively through a 100μm and a 40μm filters to generate single cell suspensions, and splenocytes were treated with red blood cells lysis buffer. Cells were stained 30 min with zombie fixable viability kit (Biolegend) in order to differentiate live cells from dead cells. Cells were further pre-incubated with Fc-Block for 15 min at 4°c and stained for 1h at 4°c with antibodies to surface markers. For intracellular stainings, cells were fixed and permeabilized with a commercially available fixation/permeabilization buffer. Cells were separated and stained using 2 panels. Extracellular staining was performed in the first panel with PerCP-conjugated CD45 (clone 30-F11), APC-H7-conjugated CD4 (clone GK1-5), BV711-conjugated CD3 (clone 17A2), FITC-conjugated CD69 (cloneH1.2F3), BV421-conjugated CD62L (clone MEL-14), BV786-conjugated CD44 (clone IM7), V500-conjugated CD8 (clone 53-6.7), BV650-conjugated CD25 (clone PC61), and in the second panel with PerCP-conjugated CD45 (clone 30-F11), APC-H7-conjugated CD4 (clone GK1-5), BV711-conjugated CD3 (clone 17A2),

BV786-conjugated CD44 (clone IM7), V500-conjugated CD8 (clone 53-6.7) and BV650-conjugated CD25 (clone PC61). Intracellular staining was performed in the first panel with APC-conjugated Foxp3 (clone FJK-16s), AF700-conjugated Gata3 (clone TWAJ), PE-conjugated RorɣT (clone Q31-378), PE-texas red-conjugated Tbet (clone O4-46), and in the second panel with APC-conjugated Foxp3 (clone FJK-16s), AF700-conjugated Gata3 (clone TWAJ), PE-conjugated RorɣT (clone Q31-378), PE-texas red-conjugated Tbet (clone O4-46), FITC-conjugated IFNɣ (clone XMG1.2), BV421-conjugated IL17A (clone TCII-18H10). Samples were analysed on a BD LSRFortessa Cell Analyzer and data were analysed using FlowJo v10.8 software.
