## Supplementary Results for "Ulcerative Colitis Host-Microbiome Response to Hyperbaric Oxygen Therapy"

**Supplementary Table 1: RNA and Protein Sequencing Panel for Digital Spatial Profiling**

| RNA Sequencing Panel | | | | | | | | |
|---|---|---|---|---|---|---|---|---|
| BATF3 | CMKLR1 | SDHA | BCL2 | CD86 | ITGB2 | VSIR | CSF1R | CXCL10 |
| TIGIT | UBB | FAS | CD276 | VEGFA | PDCD1LG2 | PTEN | CXCL9 | IL15 |
| ICOSLG | ITGAM | CD47 | MS4A1 | TBX21 | STAT3 | EPCAM | CD274 | CD3E |
| IFNAR1 | CCL5 | TNFRSF9 | HAVCR2 | HLA-E | STAT1 | PSMB10 | POLR2A | pan-melanocyte |
| CCND1 | NKG7 | PECAM1 | ICAM1 | HLA-DQ | STAT2 | CD74 | CXCR6 | CTNNB1 |
| PDCD1 | ITGB8 | ITGAX | RAB7A | IFNG | IL6 | TNF | KRT | IFNGR1 |
| HIF1A | OAZ1 | MKI67 | CD8A | IL12B | B2M | DKK2 | CD40LG | |
| **Protein Sequencing Panel** | | | | | | | | |
| CD4 | Histone H3 | SMA | HLA-DR | Ms IgG2a | CD20 | S6 | Rb IgG | CTLA4 |
| Ki-67 | Fibronectin | MsIgG1 | CD3 | CD68 | GZMB | CD8 | panCk | CD11c |
| CD56 | GAPHD | PD-1 | CD45 | PD-L1 | Beta-2 microglobulin | PD-L2 | CD127 | ICOS |
| CD80 | CD25 | CD40 | CD27 | CD44 | CD34 | FAP-alpha | CD66b | CD45RO |
| FOXP3 | CD14 | CD14 | CD163 | BIM | PARP | GZMA | Cleaved caspase 9 | BCL6 |
| BAD | NF1 | BCLXL | P53 | CD95 | Phos-Tuberin | Phos-GSK3a | PLCG1 | Phos-GSK3B |
| Phos-AKT | INPP4B | MET | Phos-AKT1 | Phos-PRAS40 | Pan-AKT | Phos-ERK | Phos-JNK | Phos-p38 MAPK |
| Phos-RSK | Pan-RAS | MAPK ERK | BRAF | EGFR | Phos-MEK1 | Phos-c-RAF | 4-1BB | Tim-3 |
| B7-H3 | STING | LAG3 | IDO1` | ARG1 | GITR | OX40L | VISTA | |

**Supplementary Table 2**: **Demographics of ulcerative colitis patients included**

| | Phase 2A trial: HBOT or sham treated UC (n=13) | Phase 2B trial: HBOT Treated UC (n=20) | Disease severity matched UC without HBOT (n=16) |
|---|---|---|---|
| Age, mean years (SD) | 44 (19) | 37 (15) | 43 (16) |
| Male gender, n (%) | 6 (46%) | 10 (50%) | 13 (81%) |
| Prior anti-TNF, n (%) | 7 (53%) | 15 (75%) | 10 (63%) |
| Prior Vedolizumab, n (%) | - | 7 (35%) | 6 (38%) |
| Prior Tofacitinib, n (%) | - | 5 (25%) | 1 (6%) |
| Mayo endoscopic sub-score of 3, n (%) | 8 (62%) | 17 (85%) | 7 (43%) |
| CRP, median (IQR) | 93 (15-123) | 14.4 (3-51) | - |
| Albumin, median (IQR) | 3.3 (3.1-3.6) | 3.5 (3.1-3.6) | - |

HBOT: hyperbaric oxygen therapy; UC: ulcerative colitis; Anti-TNF: anti-tumor necrosis factor; CRP: C-reactive protein

**Supplementary Table 3: Demographics for digital spatial profiling ulcerative colitis patients**

| Case | Baseline Endoscopy | Description | Follow-up Endoscopy and Histology | Clinical Outcome |
|---|---|---|---|---|
| **UC Patients Treated with Hyperbaric Oxygen During Hospitalization for Acute Severe Flare** | | | | |
| **1** | MES 3 with ulcers | Male, biologic and immunomodulator naïve prior to hospitalization, clinical response to HBOT by days 3 and 5 | MES 1, no friability; histology with persistence of neutrophils | Relapse requiring infliximab in 3 months |
| **2** | MES 3 with ulcers | Female, 1 prior anti-TNF agent (adalimumab) which patient was failing at time of hospitalization, clinical response to HBOT by days 3 and 5 | MES 0, normal vascular pattern; histology with persistence of neutrophils | Able to maintain response for 3 months with continued adalimumab, later relapsed requiring vedolizumab |
| **3** | MES 3 with ulcers | Male, Prior failure of 2 anti-TNF agents (infliximab, adalimumab), vedolizumab, and failing 10mg PO BID tofacitinib at time of hospitalization, clinical response to HBOT by days 3 and 5 | MES 1, no friability; histology with persistence of neutrophils | Able to remain colectomy free on 5mg PO BID tofacitinib |
| **UC Patients Treated with Standard of Care in Outpatient Setting for Acute Severe Flare** | | | | |
| **4** | MES 3 with ulcers | Female, biologic and immunomodulator naïve, started on vedolizumab after baseline endoscopy | MES 1, no friability; histology with persistence of neutrophils | Good clinical response with later requirement for Q4 week Vedolizumab |
| **5** | MES 3 with ulcers | Female, prior failure of 2 anti-TNF agents (infliximab, adalimumab), and failing vedolizumab at time of baseline endoscopy, started on golimumab after baseline endoscopy | MES 1, no friability; histology with persistence of neutrophils | Good clinical response with maintenance on golimumab |
| **6** | MES 3 with ulcers | Female, 1 prior anti-TNF agent (infliximab) which patient was failing at time of baseline endoscopy, started vedolizumab after baseline endoscopy | MES 0, normal vascular pattern; histology with persistence of neutrophils | Good clinical response with maintenance on vedolizumab |

MES: Mayo endoscopic sub-score; HBOT: hyperbaric oxygen therapy; anti-TNF: anti-tumor necrosis factor antagonist; PO: oral; BID: twice daily; Q4: every 4;

**Supplementary Figure 1: Hyperbaric Oxygen Effects on Hypoxia Response Pathways**

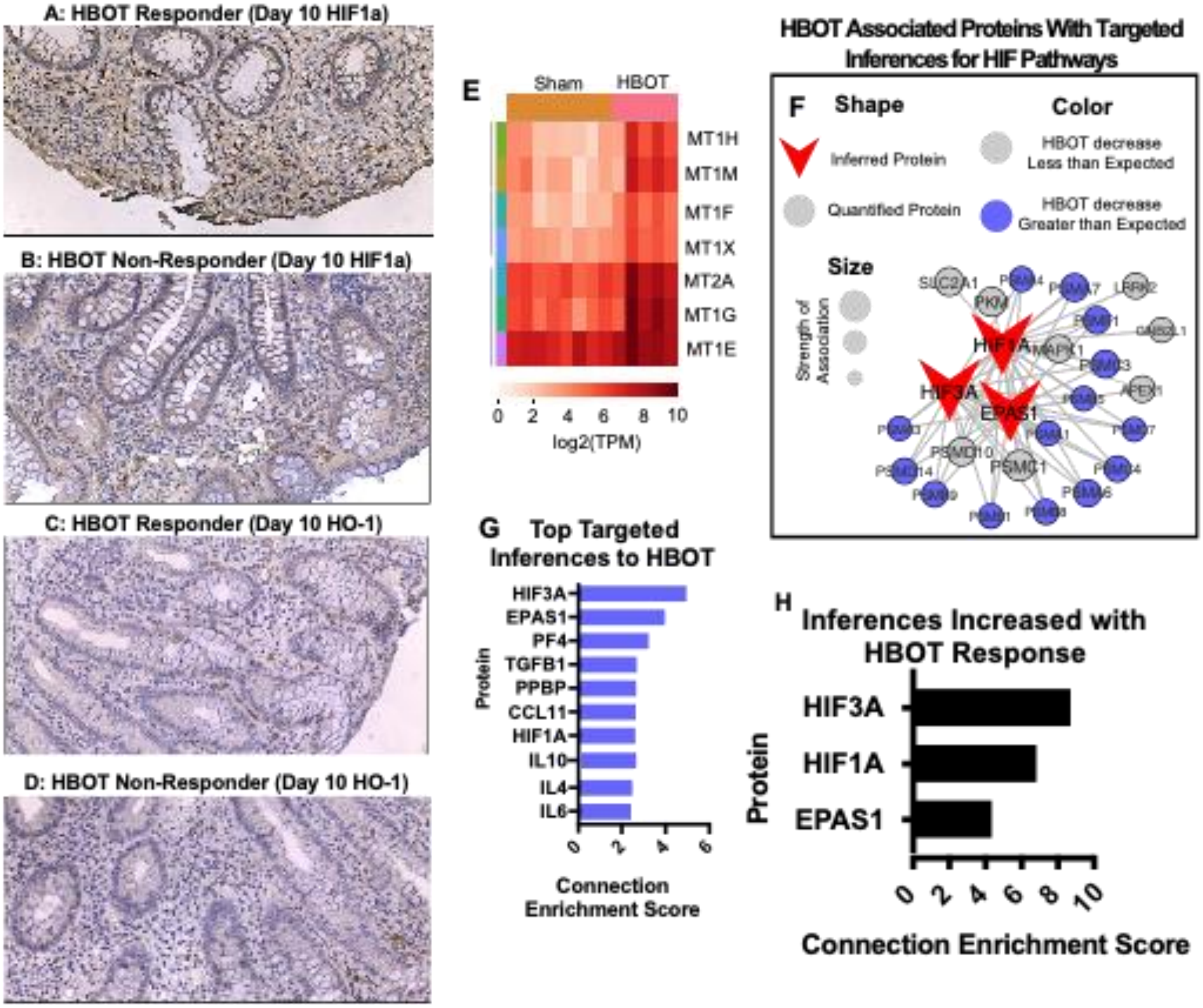


**A-D:** Immunohistochemistry staining of FFPE slides from colon mucosa on day 10 of phase 2A sham-controlled trial demonstrating comparable staining for HIF-1α and HO-1 between a HBOT responder and a non-responder. **E:** Bulk-RNA sequencing from phase 2A sham-controlled trial demonstrates a significant difference in metallothionein family members gene expression on day 10 between HBOT and sham treated UC patients (red indicates increased gene expression). All significantly increased genes (HBOT treated group versus Sham treatment control on day 10) represented as log2(TPM) normalized counts. Metallothionein family members: MT1H, MT1M, MT1G, MT1F, MT1E, MT1X; MT2A. **F:** Enrichment network graph generated using inference-based protein-protein interaction networks. Proteins labeled in red are inferred based on strength of proteins identified in the proteomics feature set, while other proteins directly observed in the proteomics feature set. **G:** Top network-inferred proteins based on all network connectivity in S1 1F, regardless of abundance directionality. **H:** Top 3 network-inferred proteins coinciding with proteins increased by HBOT treatment. Abbreviations: FFPE: Formalin fixed paraffin embedded; HBOT: hyperbaric oxygen therapy; HIF: hypoxia inducible factor 1α; HO-1: heme-oxygenase 1; MT: metallothionein; EPAS1: HIF-2α; TGFB1: transforming growth factor beta 1; PPBP: chemokine (C-X-C motif) ligand (CXCL7); CCL11: C-C motif chemokine 11; IL: interleukin; TNF: tumor necrosis factor; VEGF: vascular endothelial growth factor; CXCL: C-X-C motif ligand.

**Supplementary Figure 2: Bulk-RNA sequencing from phase 2A sham-controlled trial**

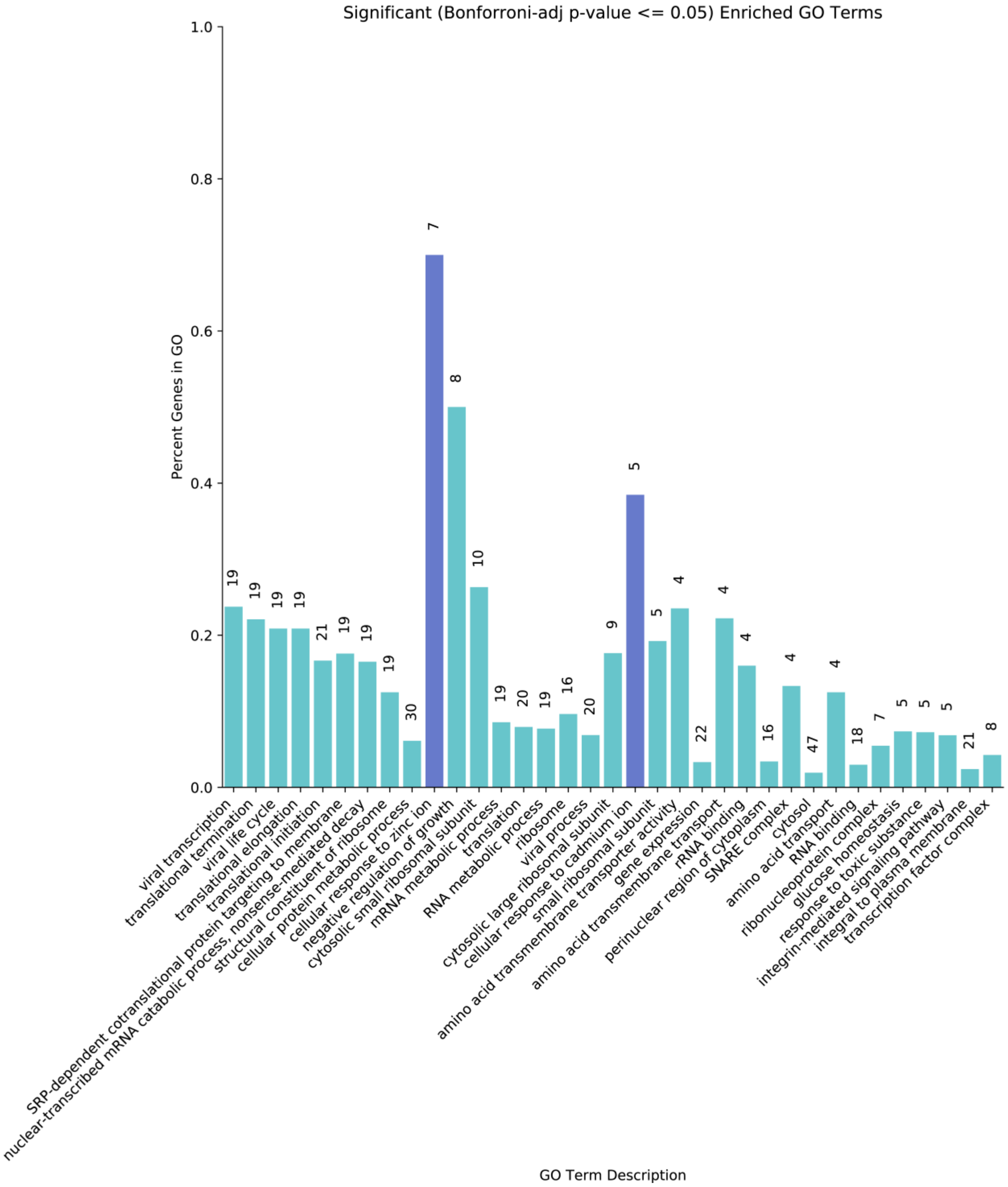


Bar graph of the percent of significantly dysregulated genes over the total number of genes within each ontological term for all significantly enriched (p-adj <= 0.05) GO terms. Blue-highlighted bars represent a focus on two terms "Cellular response to zinc ion" and "Cellular response to cadmium ion." Numbers above each bar represent the total number of significantly dysregulated genes included in each term.

**Supplementary Figure 3: Hyperbaric oxygen effect on mucosal proteins demonstrates significant effects for prostaglandin E2 synthase**

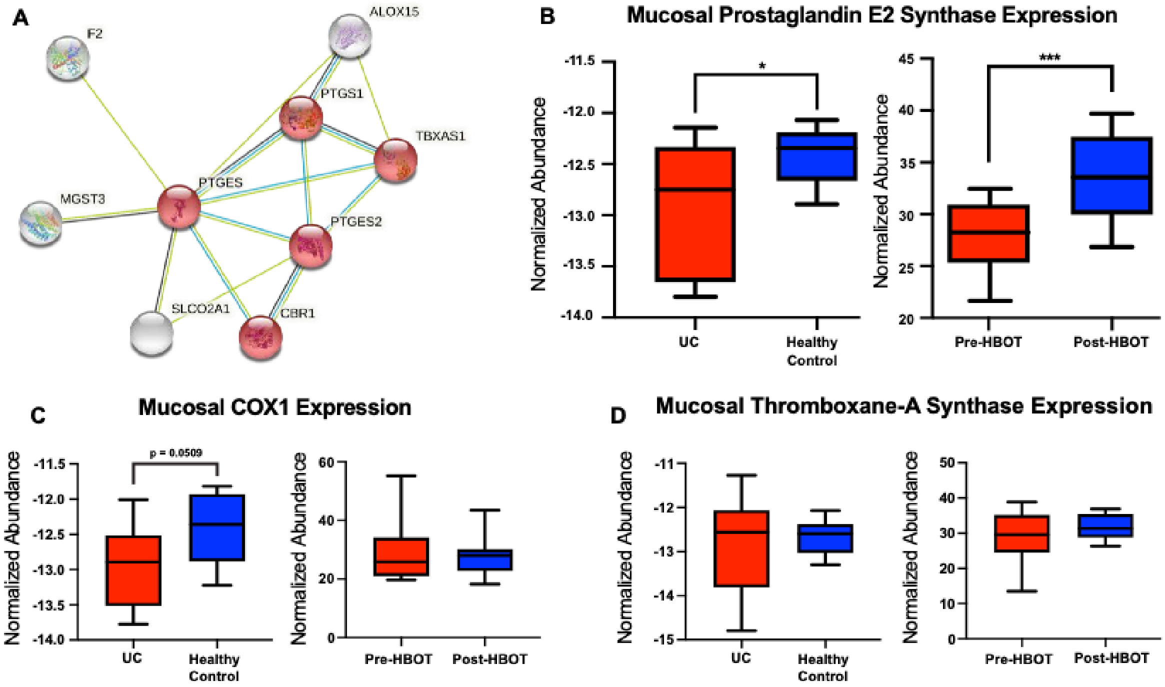


**A:** Protein-protein interaction network map of mucosal TMT-multiplexed proteomics for COX1 pathway. F2: Prothrombin; MGST3: Microsomal glutathione S-transferase 3; SLCO2A1: Solute carrier organic anion transporter family member 2A1; PTGES: Prostaglandin E synthase; PTGS1: Cyclooxygenase 1 (COX1); ALOX15: Arachidonate 15-lioxygenase; TBXAS1: Thromboxane A synthase; PTGES2: Prostaglandin E2 synthase; CBR1: Carbonyl reductase 1. **B:** Comparison of normalized protein abundance of prostaglandin E2 synthase from mucosal biopsies of UC and healthy controls (left panel) and before and after HBOT (right panel). **C:** Comparison of normalized protein abundance of COX1 from mucosal biopsies of UC and healthy controls (left panel) and before and after HBOT (right panel). **D:** Comparison of normalized protein abundance of thromboxane A synthase from mucosal biopsies of UC and healthy controls (left panel) and before and after HBOT (right panel). *p<0.05; ***p<0.001

**Supplementary Figure 4: Hyperbaric oxygen effect on fecal proteins demonstrates significant effects for azurophilic granules and NLRP3 inflammasome**

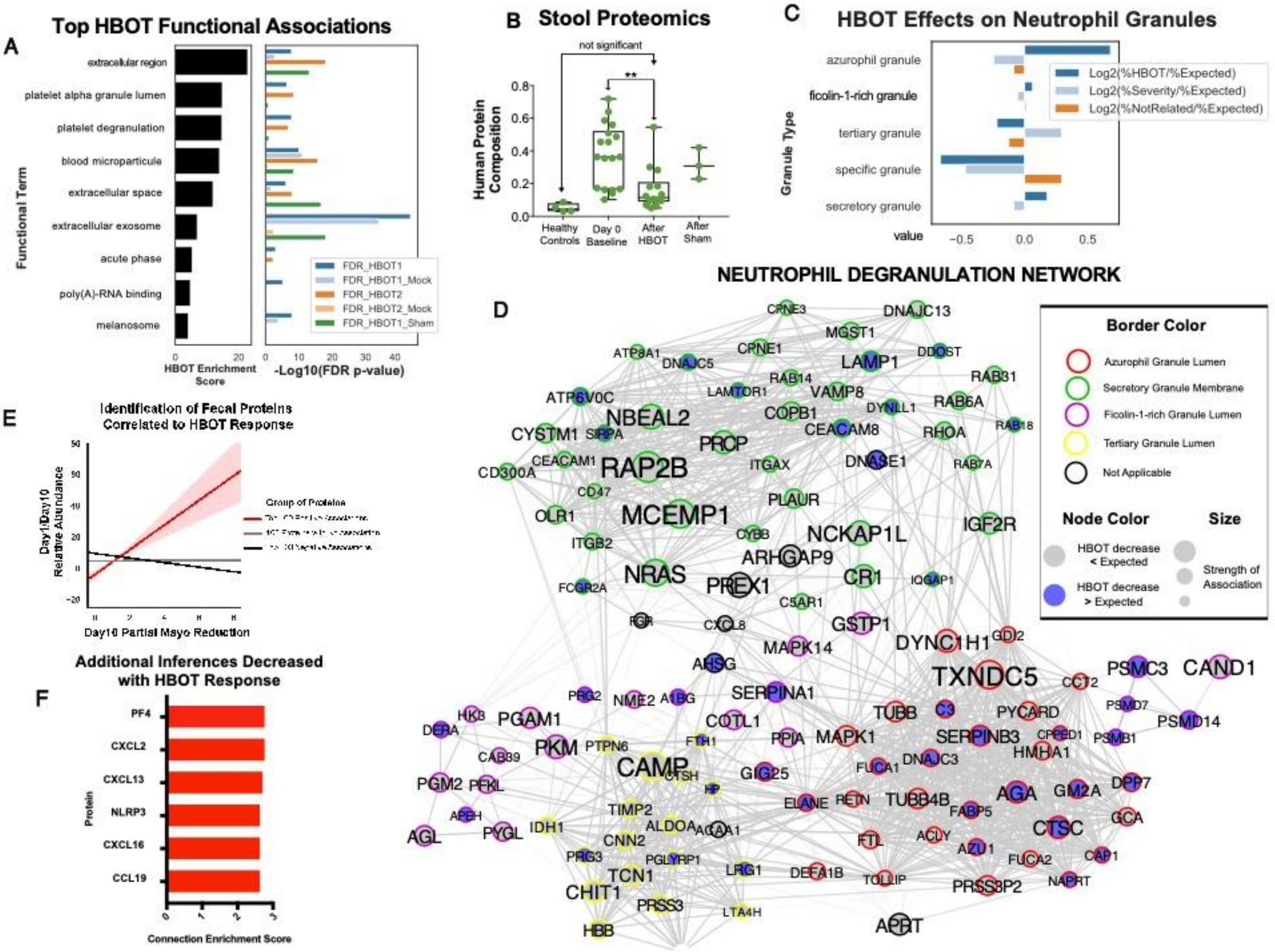


**A:** Gene ontology function terms associated with HBOT treatment (left) along with the associated FDR-generated p-value per group (right). Graph does not depict directionality of association, only strength of association **B:** Percentage of stool proteome belonging to human proteins for different groups. Significance determined by one-way ANOVA. **C:** Ratiometric comparison of neutrophil granule-specific gene ontology terms and associated enrichment value (scaled). **D:** Neutrophil-associated protein network generated by protein-protein interaction engine STRING (reformatted in CytoScape), segregated by subnetworks of granule type-specific proteins. **E:** Pearson correlational analysis of Day1/Day10 protein abundance ratios and their corresponding reduction of Partial Mayo score on Day 10. Pink area depicts the calculated confidence interval. **F:** Extended list of proteins inferred by a surrounding network of identified proteins in fecal proteome in response to HBOT treatment and their connectivity enrichment score. The surrounding network exhibited HBOT-responsive decreases in abundance, suggesting the inferred protein also decreased.

**Supplementary Figure 5: Heatmap of significantly different genes at baseline between severe UC patients requiring hospitalization and treated with hyperbaric oxygen therapy in the clinical trial, and severe UC patients able to be managed with biologics in the outpatient setting**

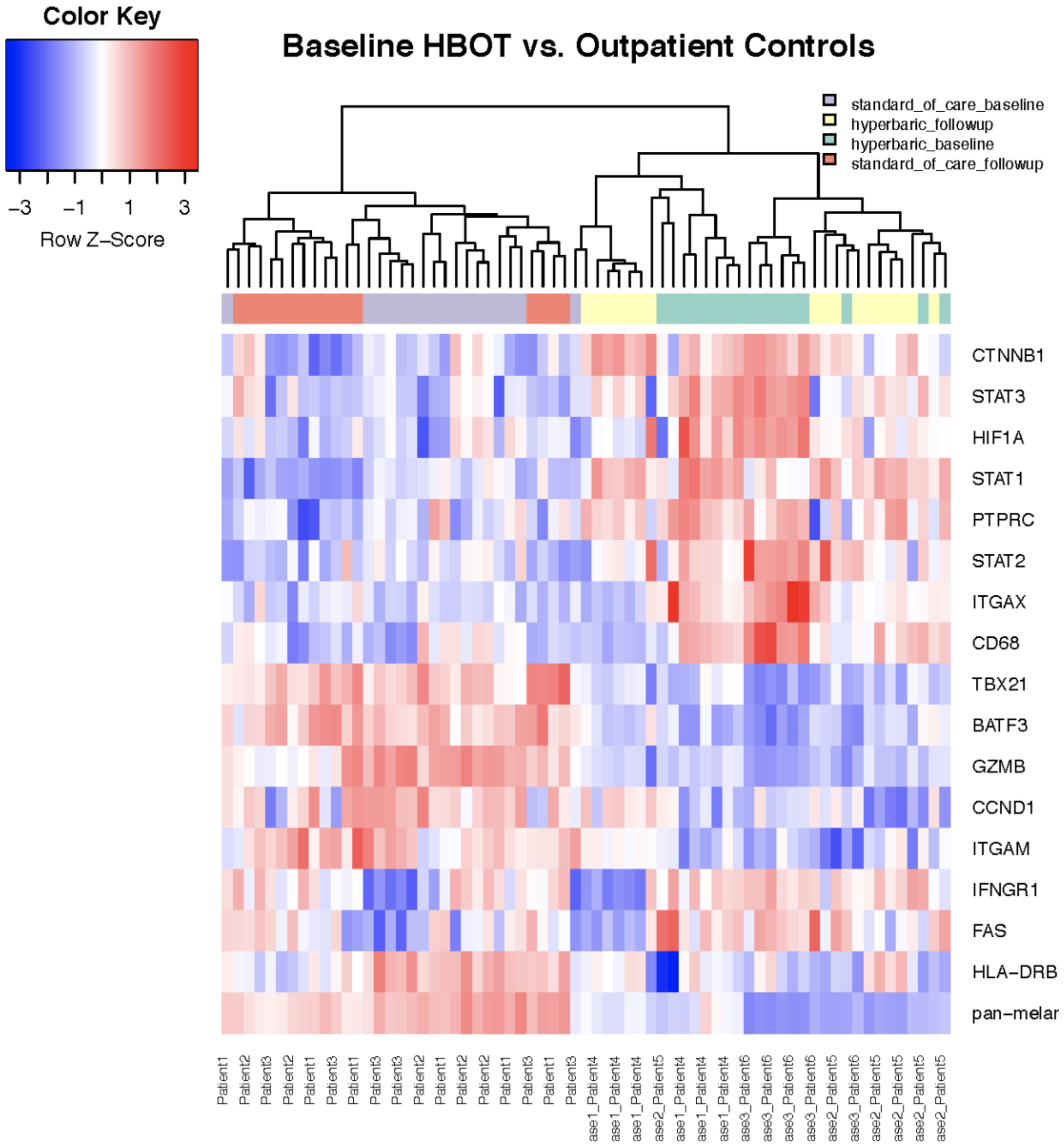

**Supplementary Figure 6: Heatmap of significantly differentially changing genes between severe UC patients requiring hospitalization and treated with hyperbaric oxygen therapy in the clinical trial, and severe UC patients able to be managed with biologics in the outpatient setting**

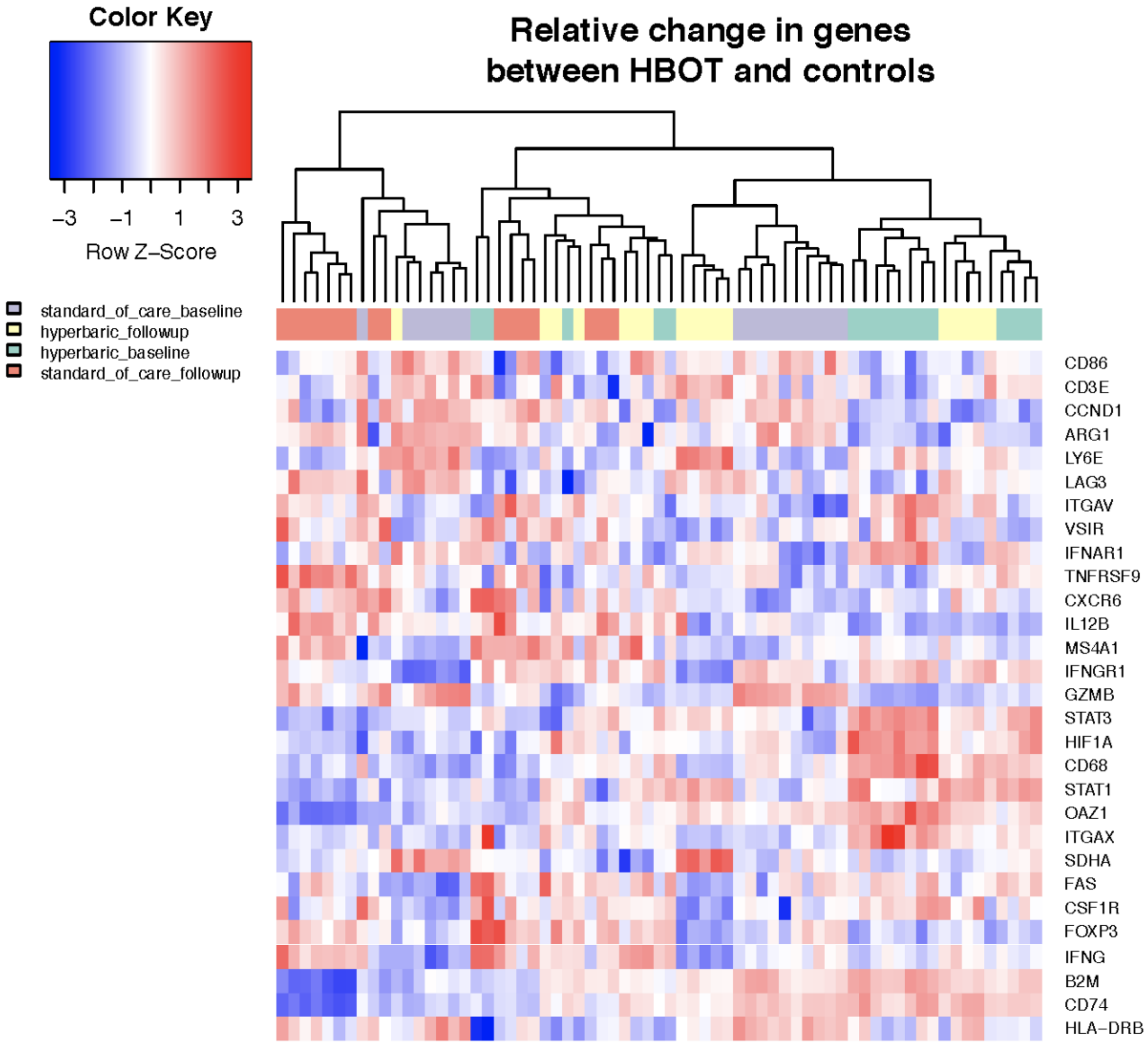

**Supplementary Figure 7: Neutrophil Gene Expression Changes in Hyperbaric Oxygen Treated Patients**

#### Pre and Post Hyperbaric Oxygen Sequencing of Neutrophil ROIs

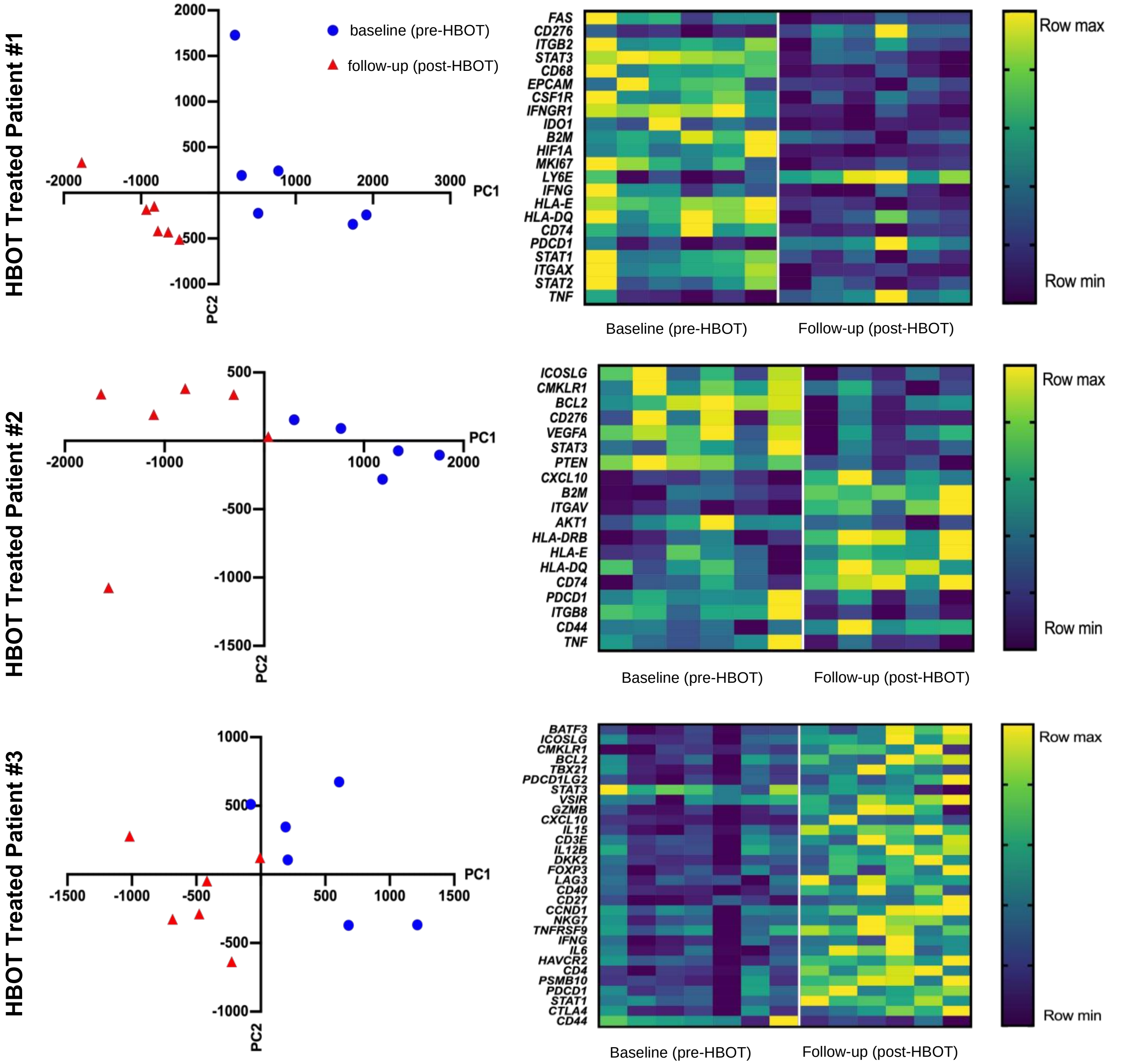

### Supplementary Figure 8: Neutrophil Gene Expression Changes for STAT3 Pathway Related Genes

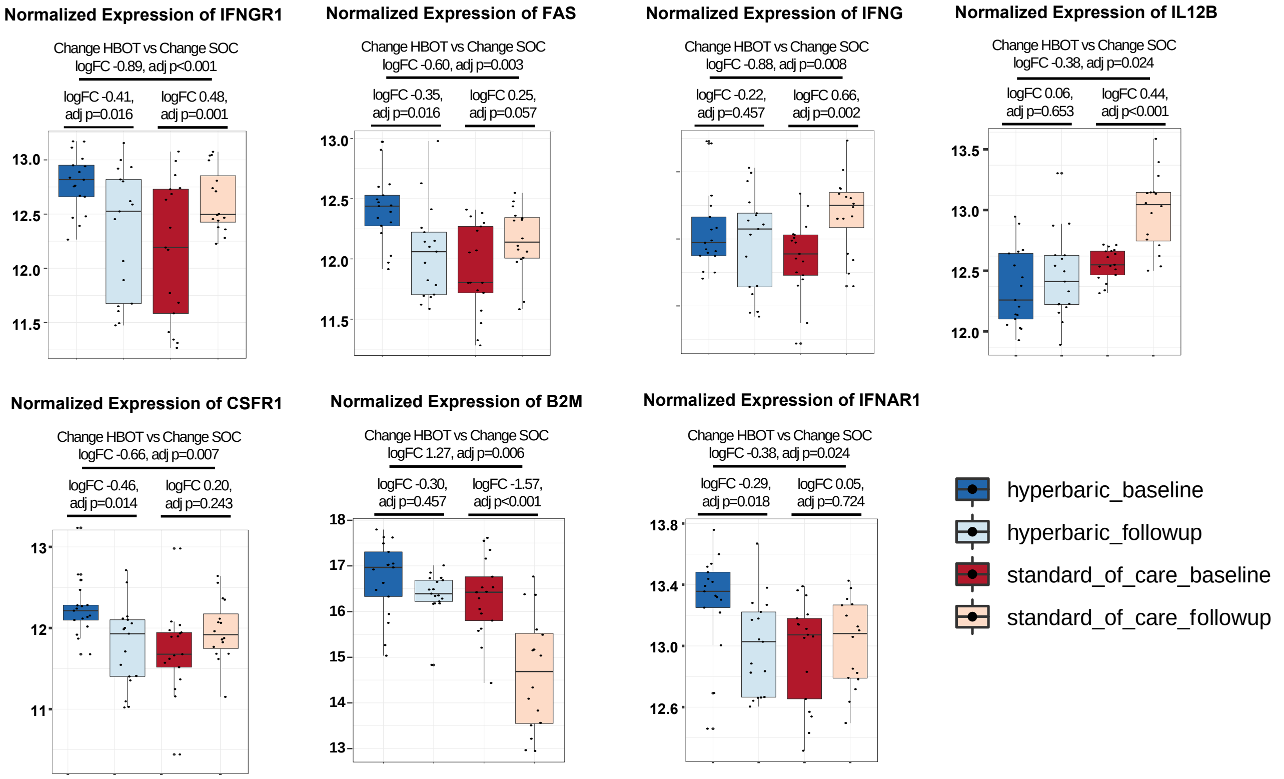

**Supplementary Figure 9: Gene Expression Changes for STAT3, STAT1, HIF1a, for neutrophils compared to other immune cell regions of interest in digital spatial profiling transcriptome**

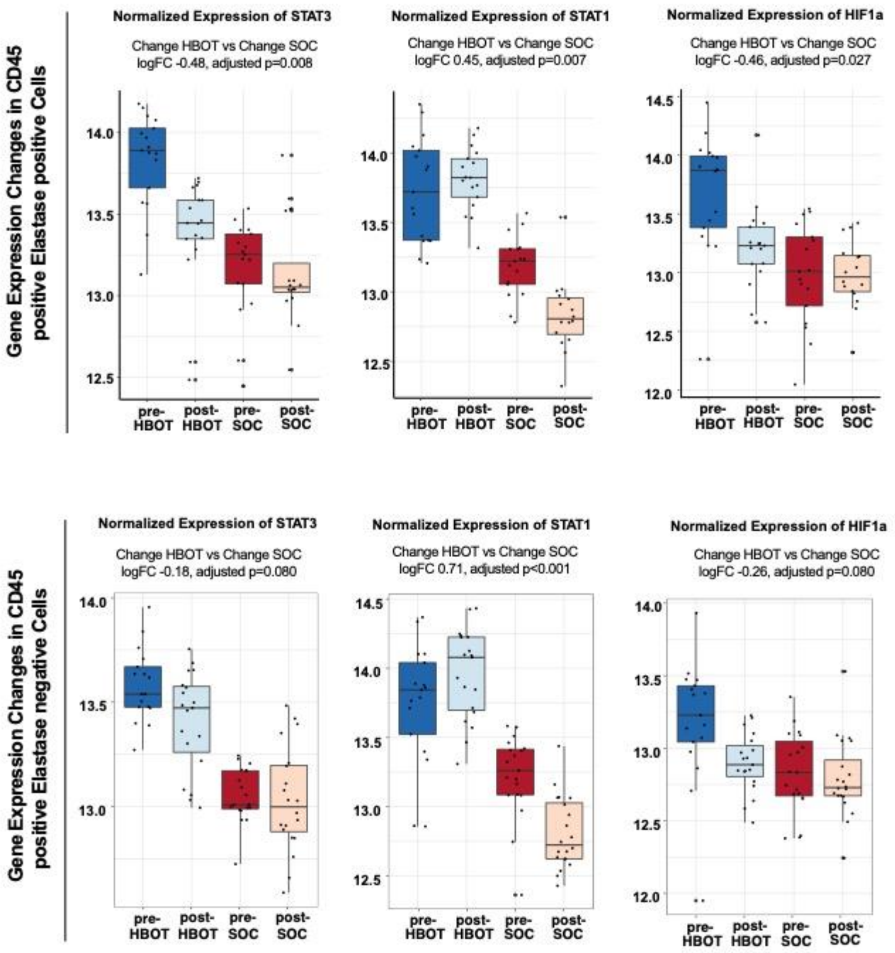

**Supplementary Figure 10: Hyperbaric Oxygen Decreases MAPK Activity with Specificity of Effect for JNK in Neutrophils**

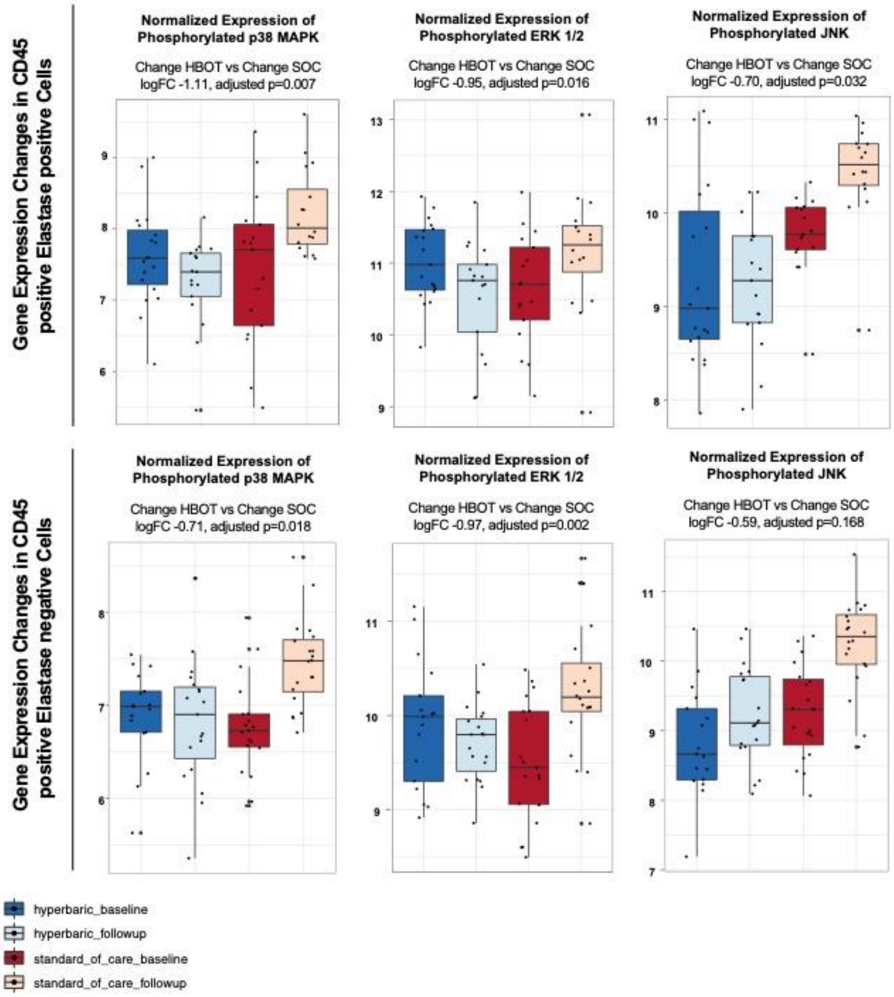

**Supplementary Figure 11: Change in diversity with hyperbaric oxygen therapy**

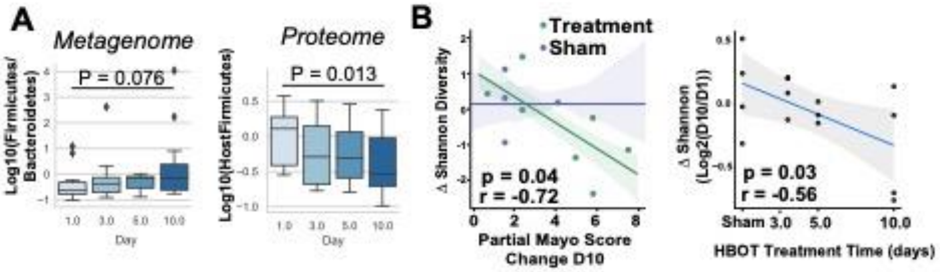

**Supplementary Figure 12: Association between HBOT response status and lithocholic acid or other metabolites**

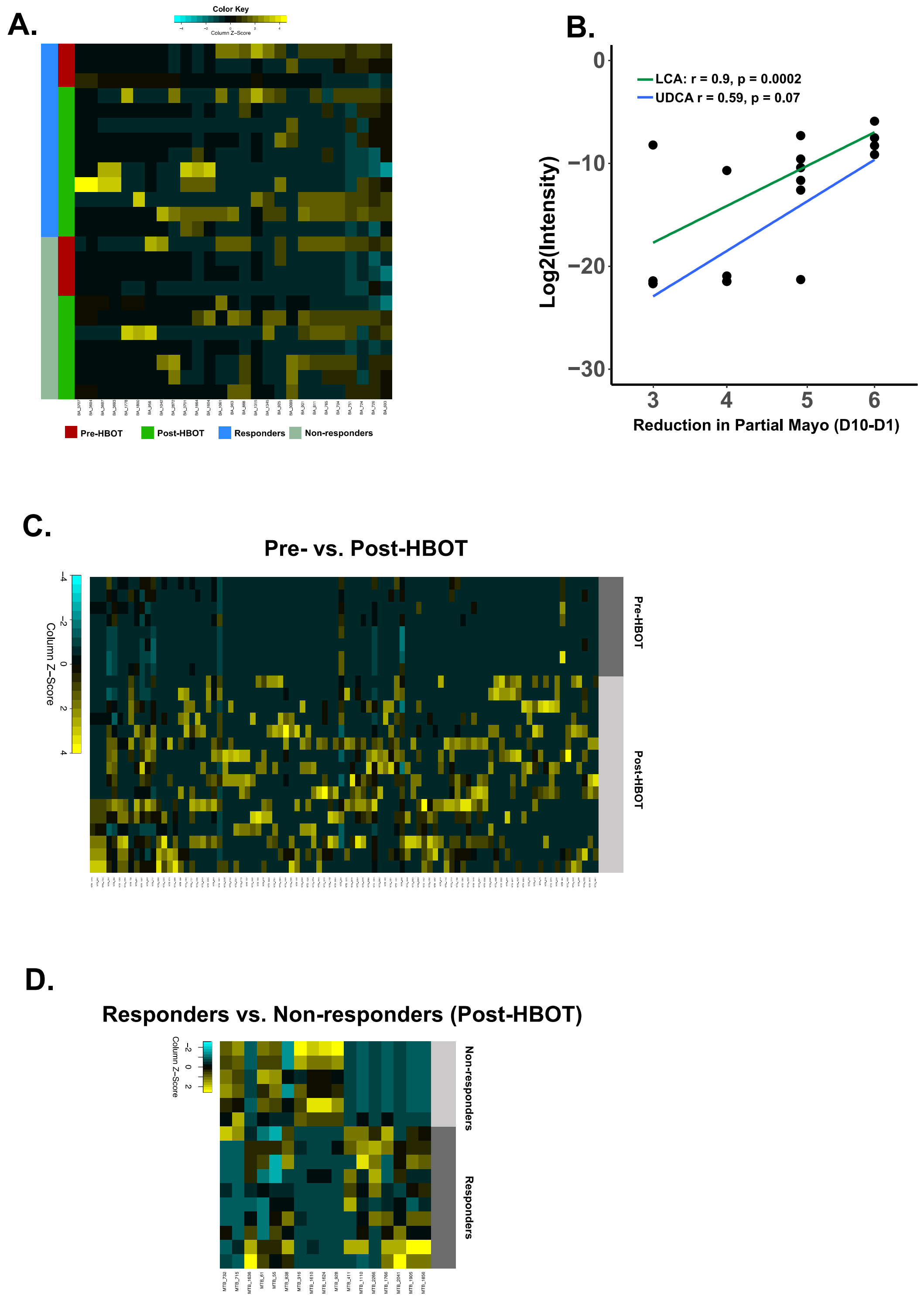


**A:** Heat map of significantly different bile acids pre- and post-HBOT according to response status**; B:** Correlation between lithocholic acid and ursodeoxycholic acid and HBOT response status**; C:** Heat map of significantly different metabolites pre and post-HBOT**; D:** Heat map of significantly different metabolites post-HBOT in responders and non-responders.

### Supplementary Figure 13: MUC protein changes in mucosal biopsies and stool with hyperbaric oxygen therapy

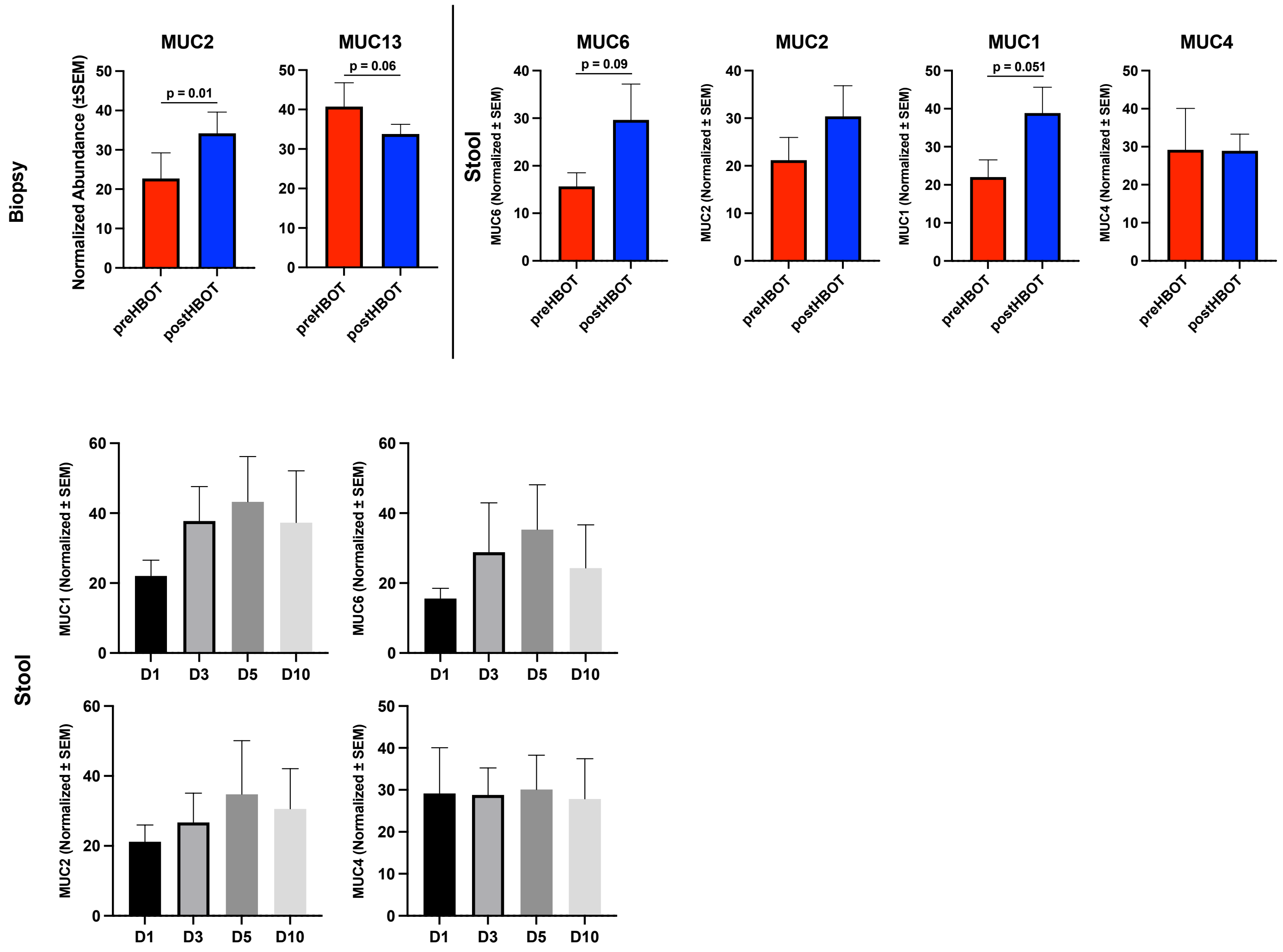

**Supplementary Figure 14: Association between changes in fecal mucus proteins and *Akkermansia muciniphila* strains**

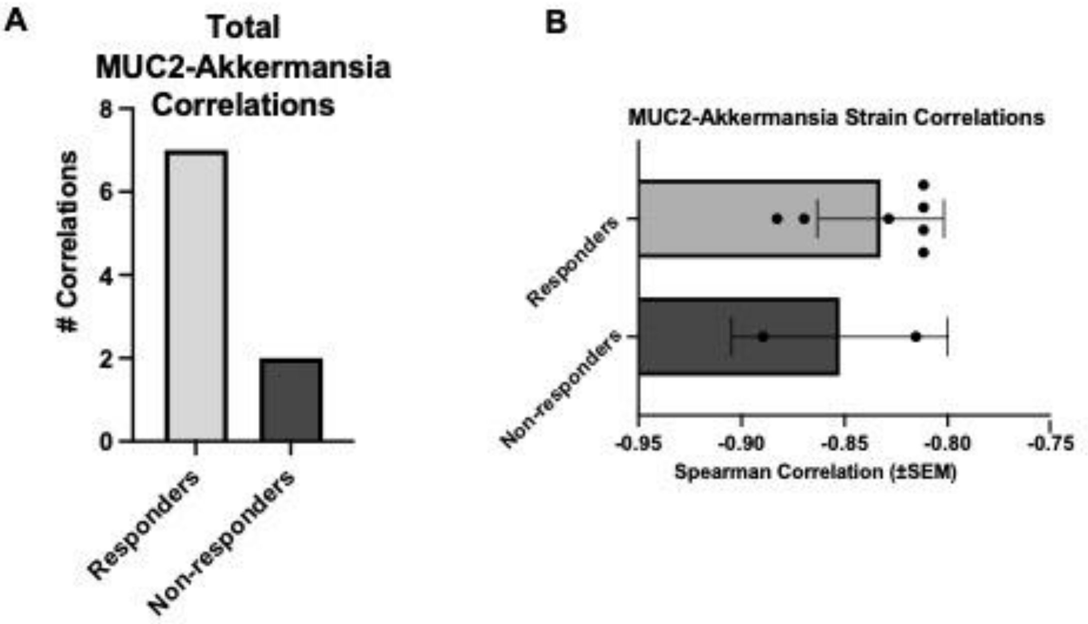


**A)** Total significant ($p < 0.05$, Spearman) correlations of stool-detected MUC2 protein levels and post-HBOT abundance of various *Akkermansia* strains identified, split by responders and non-responders. **B)** Stool-identified MUC2-levels and their correlation (Spearman) to post_HBOT *A. muciniphila* levels. Each dot represents a specific correlation between each pair.

#### Supplementary Figure 15: qPCR from mouse colonization experiments

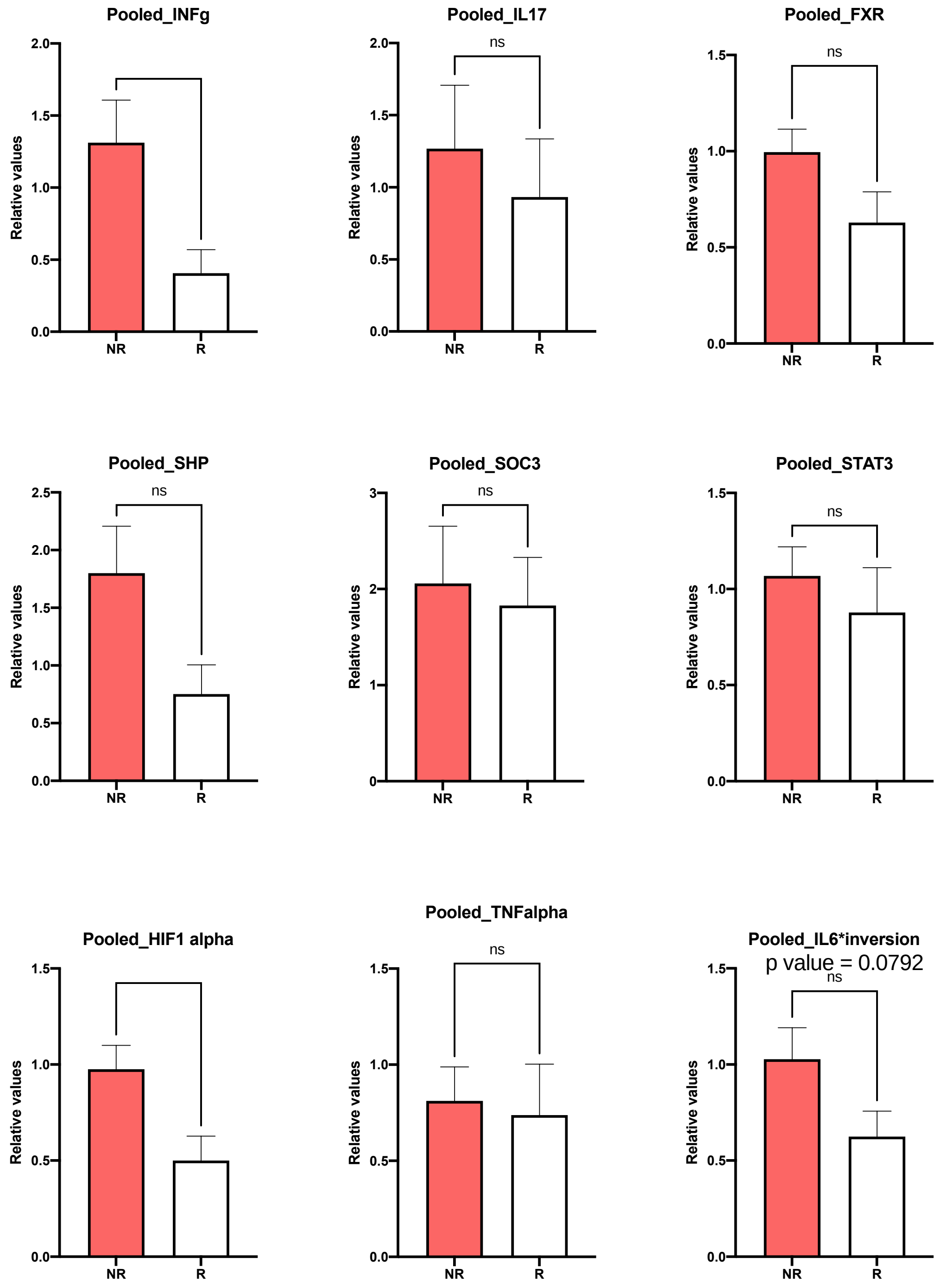
